# Out-of-hospital births and the experiences of emergency ambulance clinicians and birthing parents: A scoping review of the literature

**DOI:** 10.1101/2024.11.09.24316932

**Authors:** Michella G. Hill, Alecka Miles, Belinda Flanagan, Sara Hansen, Brennen Mills, Luke Hopper

**Author notes:** Corresponding Author Michella Hill, Edith Cowan University, School of Medical and Health Sciences 270 Joondalup Drive, Joondalup Perth, Western Australia 6027, Facsimile.

## Abstract

**Objective:** Emergency ambulance services attend a wide array of medical and trauma patients. Infrequently, this includes imminent or out-of-hospital births (OOHBs). There is a paucity of research pertaining to OOHBs. This scoping review explores emergency ambulance clinician involvement with OOHBs, and patient and clinician experiences with birthing in the out-of-hospital setting.

**Design:** Scoping review; two reviewers independently determined inclusion using the Joanna Briggs Institute framework and ‘participant, concept, context’ criteria.

**Methods:** CINAHL, Embase, Medline, Web of Science, and Wiley Online were searched from database inception until 20 February 2024. Articles discussing an unplanned OOHB, or a planned home birth with complications where an emergency ambulance was required were included.

**Results:** Sixty-three articles were included for review. The majority (*n*=36) involved retrospective research. Most articles were published since 2015 (*n*=38), with the highest contributing countries being USA (*n*=17) and Australia (*n*=13). Risks factors for OOHBs were varied with maternal age or being multigravida/multiparous often cited. Ninety-nine complications were described in the literature ranging from relatively minor ailments such as nausea and vomiting through to life-threatening situations such as maternal or neonatal cardiac arrest. The most common management/interventions reported were assisting with birth, maternal intravenous cannulation, and medication administration.

Birth parents, partners and clinicians all describe OOHBs as anxiety-provoking but joyous when a healthy neonate is born. The OOHB experience is enhanced for patients when clinicians communicate well, while those who appeared inexperienced increased patient anxiety.

OOHBs experience many challenges to optimal care, falling under the broad categories of ‘emergency ambulance clinicians desiring additional education and training’, ‘communication and collaboration difficulties’, ‘environmental issues’, ‘technology and aids’ and ‘other’ limitations.

**Conclusion:** OOHBs are rare events requiring expert assistance to optimise patient outcomes. There remains significant challenges to unplanned OOHBs; ongoing training and skill competency is required to improve patient safety and clinician confidence.

**Article Summary:** *Strength and limitations.:*

- This review provides a comprehensive overview of unplanned OOHBs attended by emergency ambulance clinicians in high-income countries; this also includes planned home births or freebirths where emergency ambulance assistance was required.
- We utilised a rigorous methodology framework as per the JBI guidelines and the Preferred Reporting Items for Systematic Reviews and Meta-Analysis extension for Scoping Reviews.
- This review did not consider risk of bias, rigour, or quality from included studies, however most research in this space is from retrospective or qualitative research methodologies.
- Only peer-reviewed, full text publications in English were included. Educational material, conference papers, letters to the editor, or opinion articles were excluded.

*Review registration:* This scoping review is registered with the Open Science Framework (https://osf.io/bd62h), registration DOI 10.17605/OSF.IO/TA35Q.

**Summary Box:** *What is already known on this topic:* Unplanned out-of-hospital births occur rarely, yet have the capacity to be life-threatening events for both birth parent and neonate.

*What this study adds:* This study provides an extensive overview of emergency ambulance clinician involvement in unplanned out-of-hospital births, birth complications and interventions provided. Furthermore, it comprehensively explores the patient and clinician experience, which is frequently described as traumatic and anxiety-provoking, with evidence suggesting support services such as telehealth could aid both the patient and emergency ambulance clinician in these situations.

*How this study might affect research, practice, or policy:* Quality ongoing training, education, and exposure to birthing for emergency ambulance clinicians is strongly recommended. Ensuring emergency ambulance service guidelines have been appropriately adapted for unplanned out-of-hospital birth is also encouraged, as it appears some guidelines (such as resuscitative hysterotomy in maternal cardiac arrest) are unrealistic for the out-of-hospital environment.

## Introduction

Emergency ambulance service caseload varies widely, ranging from high impact trauma to low-acuity or chronic medical presentations.(1) Obstetrics care is included in this diverse workload with pregnant patients presenting with perinatal emergencies or non-obstetrics related presentations.(2) Occasionally, it can involve assisting with precipitous births or aiding with a planned home birth where the midwife has requested emergency ambulance assistance for the neonate or birth parent following unanticipated complications. Births which occur in an unplanned location in the out-of-hospital (OOH) environment are known as ‘births before arrival’ (BBA) or ‘out-of-hospital births’ (OOHBs).(2)

The incidence of intrapartum cases attended by emergency ambulance clinicians is relatively low compared to other callouts. For example, one study found intrapartum cases represented 0.05% of the Queensland Ambulance Service caseload in 2010–2011, with around 10% of these progressing to an OOHB in paramedic care. While this percentage sounds negligible, intrapartum cases amounted to 6,135 callouts.(3) The Australian Institute of Health and Welfare reports annually on Australia’s Mothers and Babies. This report advises in 2021 the recorded place of birth for 7% of women was ‘other’,(4) which includes births that did not occur in the planned location, freebirths where there was no clinician in attendance, births at community health centres, and OOHBs. ‘Other’ births appear to be increasing, up from 4% in 2016.(4) While it is unknown how many of these ‘other’ births were attended by emergency ambulance clinician’s, these rare events are often classified as ‘low-frequency, high-risk’ callouts.

Unfortunately, OOHBs have an increased incidence of adverse outcomes for both the birth parent and neonate compared to hospital, maternity unit or planned home births.(2,5) Complications may be unanticipated and unavoidable such as preterm births (defined as <37 weeks gestation), but also include postpartum haemorrhage (PPH) and neonatal hypothermia which must be effectively managed OOH to avoid adverse outcomes.(2,5) Birth parents and emergency ambulance clinicians have described OOHBs as distressing and sometimes requiring psychological support following their experiences.(6,7) Some emergency ambulance clinicians have indicated they feel underprepared to handle an OOHB or obstetrics emergencies and report a lack of recent education and training on the subject.(8) Given the infrequency of OOHBs, obstetric-specific clinical knowledge and skills may be untested for years before being required in a high-risk emergency.

It is essential that emergency ambulance clinicians maintain their specialised obstetrics skills and knowledge above the minimum competencies required to ensure the well-being of both the birth parent and neonate (or foetus). In an emergency, this knowledge must be readily retrievable for patient safety, and ideally the clinician should be confident in their abilities.

The objective of this review is to explore:

- Emergency ambulance clinician involvement in OOHBs, including incidence, risk factors and complications;
- Birth parent, significant partner, and emergency ambulance clinician experiences with OOHBs; and
- The barriers and challenges in providing optimal treatment provision for unplanned OOHBs by emergency ambulance clinicians.

## Methods

The Joanna Briggs Institute (JBI) framework and methodology for conducting scoping reviews (9–11) was used with the Preferred Reporting Items for Systematic Reviews and Meta-Analysis extension for Scoping Reviews (PRISMA-ScR) guidelines reported.(12) The protocol for this review was published a-priori which provides comprehensive details relating to the search protocol.(13)

### Eligibility criteria

Articles were included in line with the ‘population, concept, and context’ criteria recommended by Lockwood et al.(12) and search terms were devised in consultation with two Edith Cowan University librarians. For the purposes of this review, the *population* is birthing parents who have an out-of-hospital birth and the emergency ambulance clinicians who attend them. The *concept* is birth, which includes imminent births and/or immediate postpartum care provided by emergency ambulance clinicians. The *context* is out-of-hospital emergency care, and occurred in a ‘high-income’ country as defined by the World Bank.(14) Exclusions related to obstetric patients treated for non-birth-related presentations, planned home births with midwifery assistance not requiring an emergency ambulance, births in hospitals or birthing clinics, post-birth institutional transfers for birth parent and/or neonate, educational materials, textbooks, and abstracts without the full article.

### Search strategy

Utilising the three-step process recommended by JBI, an initial search of the CINAHL and Medline databases was conducted on 22–23 November 2022 following the priori.(13) Located articles were analysed for text words used in the Title and Abstract, and the index terms used to describe the publication. A second search of the databases CINAHL, Embase, Medline, Web of Science and Wiley Online was with articles included up to 20 February 2024. The full list of keywords used to conduct the full search are described below.

Population: ambulance* OR “emergency medical service*” OR “emergency medical technician*” OR aeromedical OR “helicopter transport*” AND

Concept: birth* OR “born before arrival” OR childbirth OR deliver* OR “emergency birth” OR labo*r OR matern* OR neonat* OR newborn OR obstetric* OR outborn OR parturition OR perinat* OR postpartum AND

Context: “out of hospital” OR “out-of-hospital” OR “out of institution” OR “out-of-institution” OR paramedic* OR pre*hospital

The third step involved reviewing the reference lists of selected articles to locate additional studies. To reduce selection bias and inclusion error two reviewers (MH, AM) independently screened publications to ensure the articles met the criteria.

### Data charting

A data extraction tool was utilised to logically chart key information from each included publication. Extracted information included authors, year of publication, title, country, study aims, participants, methods and results relevant to research questions. Three articles were independently assessed by two reviewers (MH, BM) and the extracted data was compared to ensure relevant data was included. Further data extraction was predominantly undertaken by one researcher (MH). The data extraction tool has been included as a Supplementary, Table 1. A quality assessment of each article was not undertaken for this review. A descriptive summary of findings has been provided.

### Ethics

This study does not require ethical review as all the information obtained comes from publicly available resources. This scoping review has been registered with Open Science Framework (https://osf.io/bd62h), registration DOI 10.17605/OSF.IO/TA35Q.

## Results

The final analysis of the five databases yielded 9,649 records for consideration and 230 duplicate records were removed. Figure 1 below shows a detailed PRISMA-ScR flow diagram(15) illustrating the identification, screening, and selection process for the 63 included articles.

**Figure 1:**
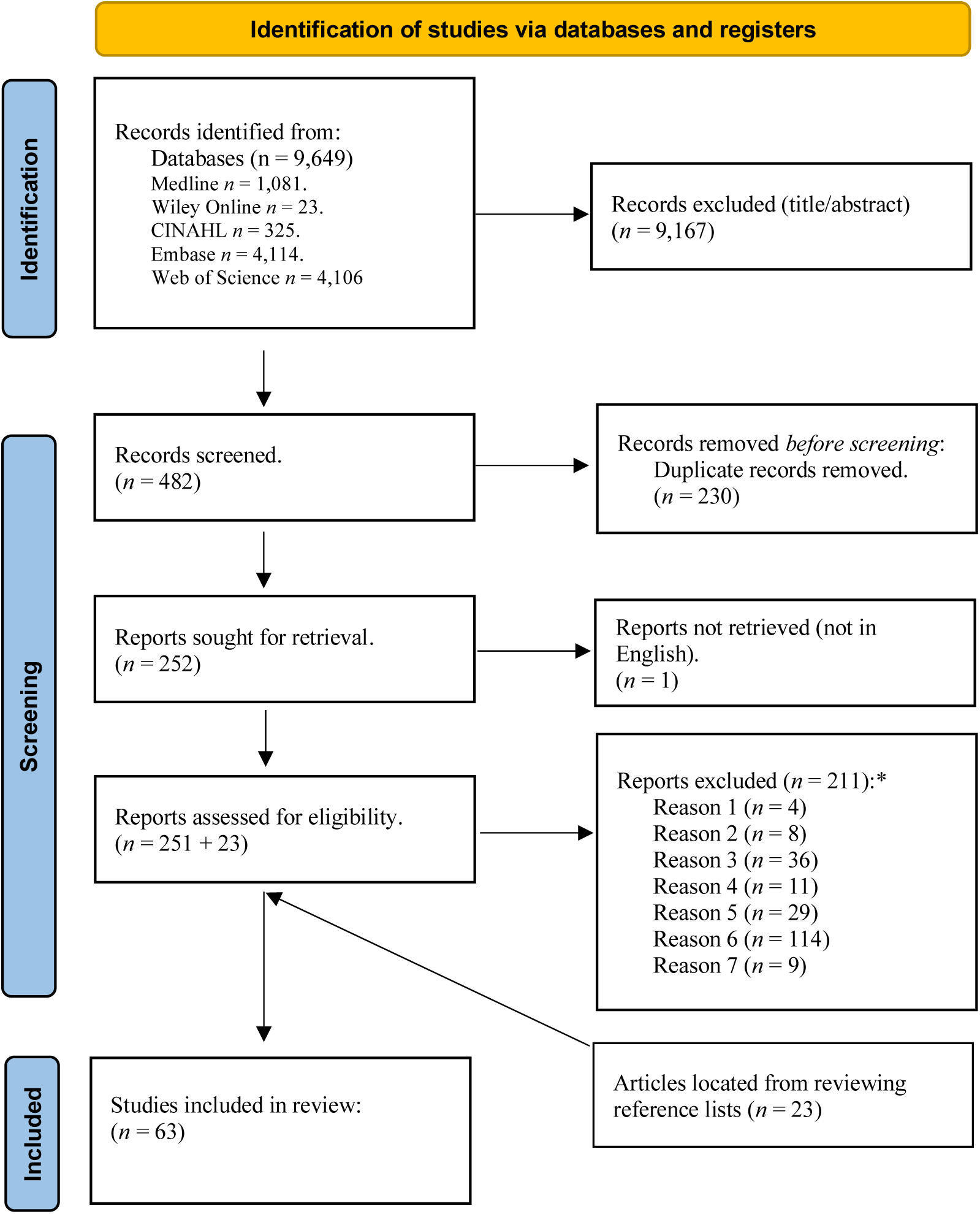
Selection process for included articles in the scoping review. *Reasons for exclusion, Reason 1 – were Review articles; removed to avoid duplication of data. (Ensured all relevant articles were already considered for this review). Reason 2 – were not in English. Reason 3 – were educational material, textbooks, abstract only, Letters to the Editor, or discussion article (not research). Reason 4 – according to World Bank, research not done on a ‘high-income’ country. Reason 5 – births under midwifery care where ambulance assistance is not required and/or maternity unit/institutional births. Reason 6 – not relevant to research questions (e.g., interfacility transfers of neonate post-birth; simulation exercises) Reason 7 – birth occurred after transportation to hospital. No automation was utilised in the screening process (all manually screened).

### General characteristics of included articles

The top three countries contributing studies to this review were the United States of America (USA) (*n*=17), Australia (*n*=13) and the United Kingdom (UK) (*n*=8). Figure 2 provides a comprehensive summary of contributing countries.

**Figure 2.**
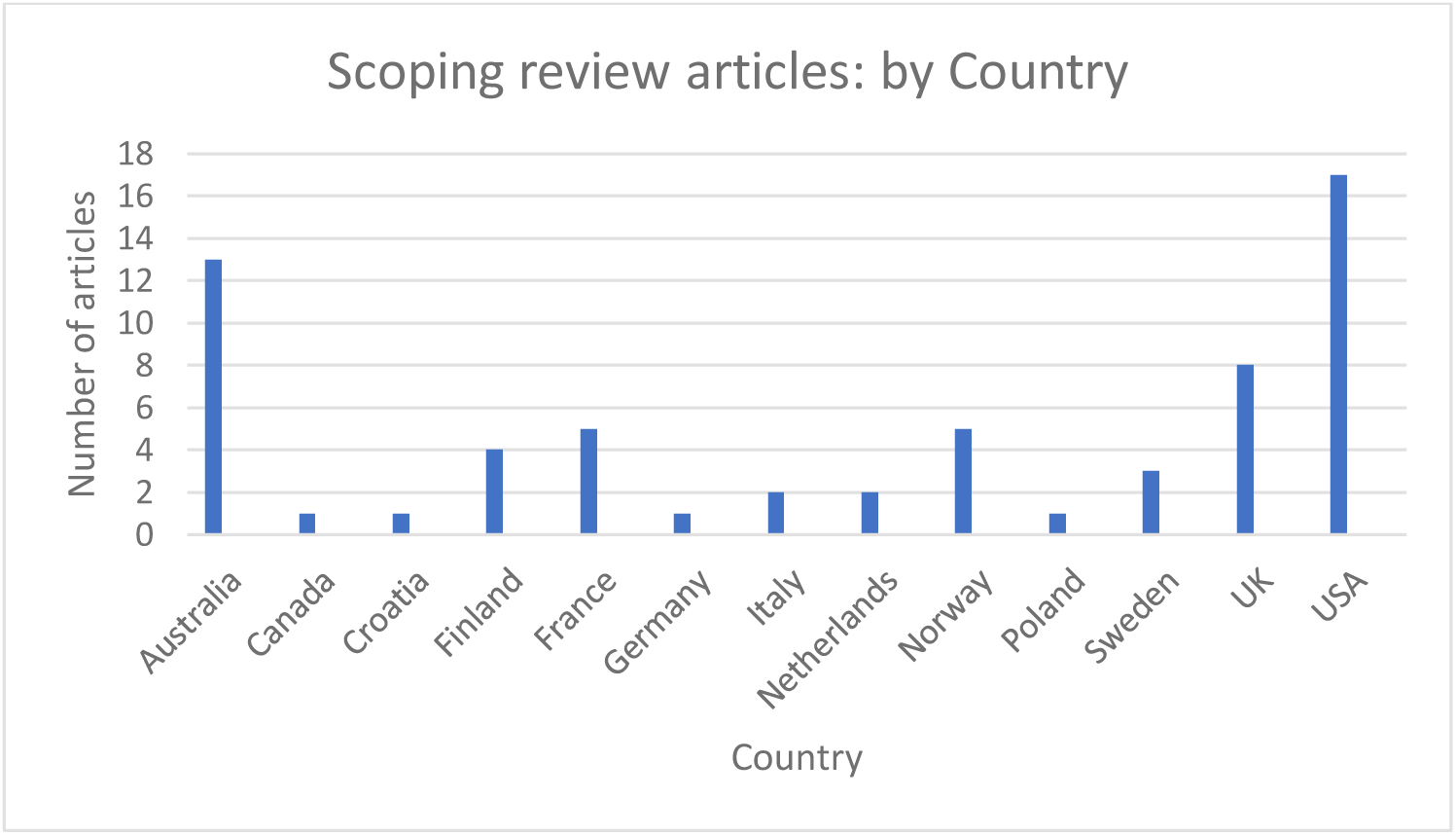
Countries which contributed articles to the scoping review

Figure 3 shows the methodologies utilised for the included articles, with retrospective data (i.e.: historical patient care records or hospital data) being the most widely used study design.

**Figure 3:**
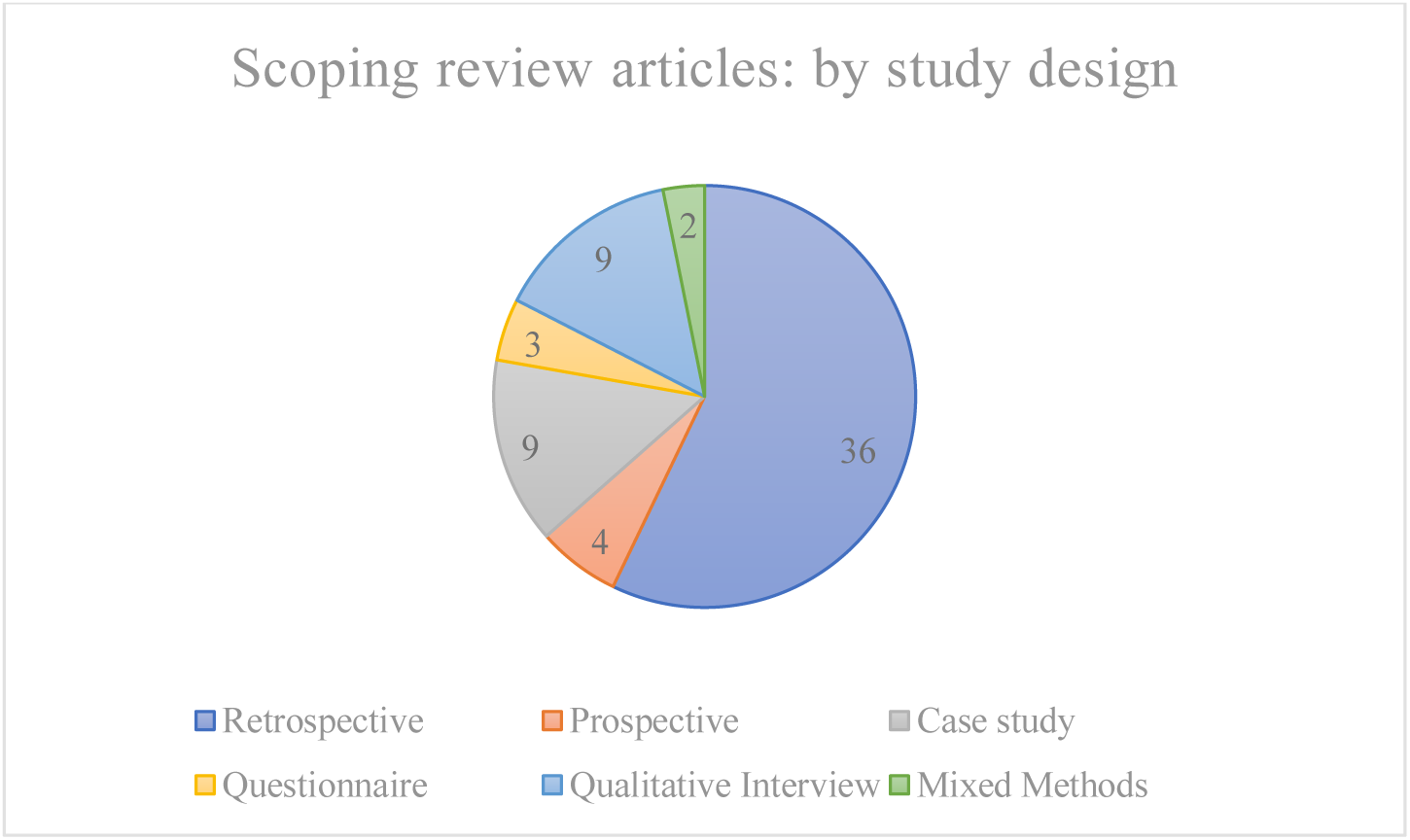
Research methodologies employed for articles included in the Scoping Review

The number of participants pooled across the research amounted to 1,868,304 (Table 1). The number of neonates born OOH amounted to 40,596. There were only 85 emergency ambulance clinicians represented in the data set. It should be noted that while home births were specifically excluded from this review, articles which discussed home births where emergency ambulance clinicians became involved in the birth were still included.

**Table 1:** Total number of participants included in the Scoping Review.

| Neonates – OOHBs | Neonates – case-controls | Planned home births | Antenatal or intrapartum transfers | Birth parents | Partners of birth parents | Emergency ambulance clinicians | Total |
| --- | --- | --- | --- | --- | --- | --- | --- |
| 40,596 | 1,671,043 | 29,600 | 126,644 | 323 | 13 | 85 | **1,868,304** |

Publication dates ranged from 1987 to 2023, with a higher frequency of publications seen since 2015 (Figure 4).

**Figure 4:**
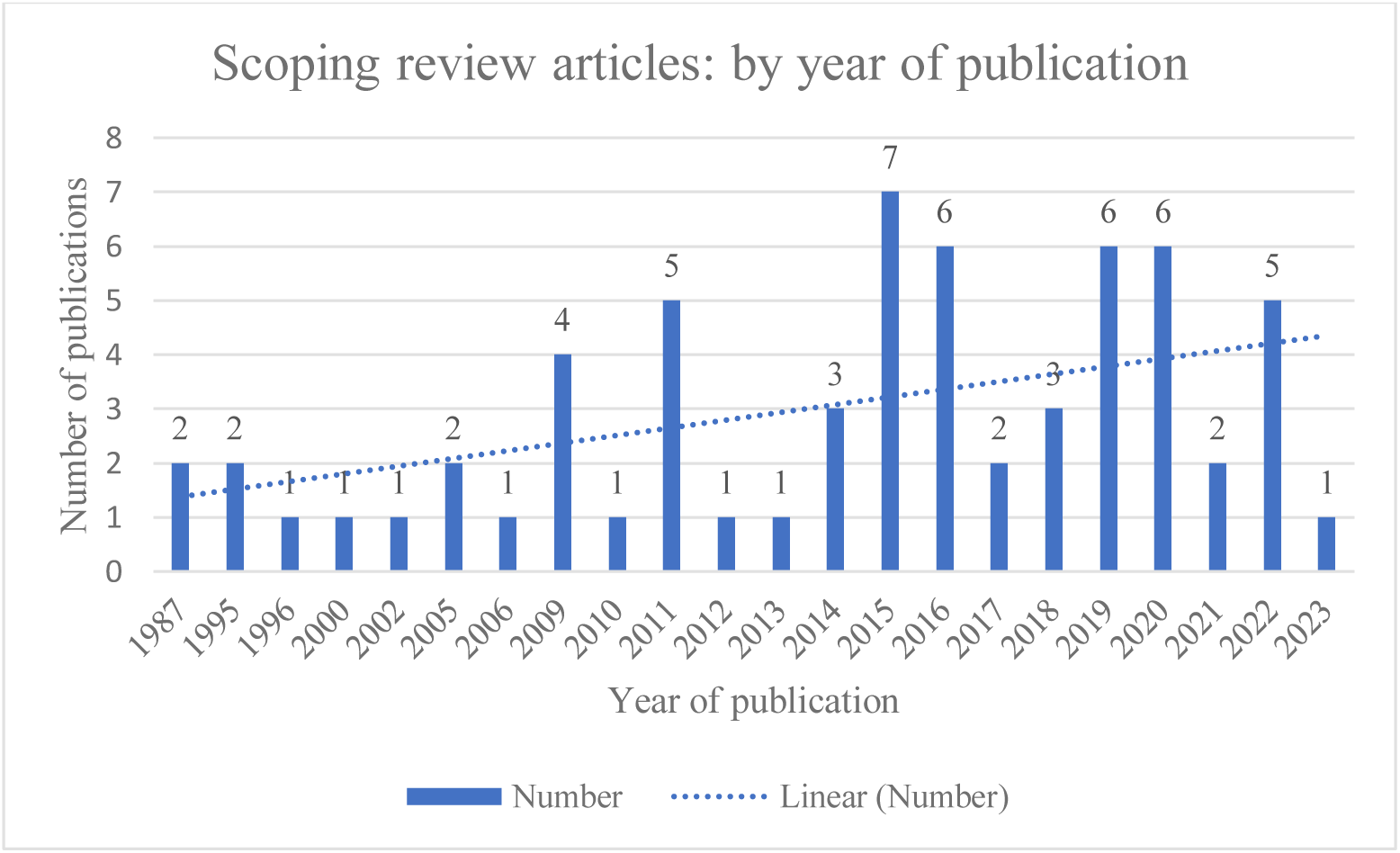
Number of included articles published by year.

### Emergency ambulance involvement in OOHBs, including incidence, risk factors and complications

#### Incidence

The incidence of imminent birth callouts for ambulance services varied from 0.5% – 0.6%,(3,16) while actual OOHBs were reported at 0.02% – 0.3%.(3,16–22) For aeromedical retrievals, one service reported neonates born within 24-hours of the call request comprised 2.7% of all paediatric cases.(23) Maternal cardiac arrests, while extremely rare, contributed a number of articles in the scoping review (*n*=4),(24–27), with many articles discussing specific case studies reporting on management and patient outcomes.

#### Risk Factors

Risk factors for OOHBs and increased neonatal morbidity and mortality are depicted in Supplementary Table 2. The analysis of risk factors was highly heterogenous between studies. Risk factors such as ‘maternal age (older)’ or ‘birth weight (low)’ were often not defined with exact integers due to differing definitions between studies. A few articles used descriptive analysis reporting on ‘known risk factors’ from patient characteristics, whilst others used proportional analysis. Some studies compared OOHBs to another cohort and used statistical significance to determine possible risk factors. Articles using methodologies such as univariate or multivariate regression modelling have a separate citation column in Supplementary Table 2, to separate these studies from the less rigorous methodologies. Two articles discussed different factors in their regression analysis and were not included in Supplementary Table 2: Akl et al.(28) reviewed aeromedical transfers for obstetrics patients, and found no in-flight births, although there were two births during ground transportation.

However, the clinical factors which were associated with shorter duration between landing and birth were nulliparity, cervical dilation ≥4cm, ruptured membranes, and a gestational age of >32 weeks. Javaudin et al.(29) assessed what factors were associated with significant improvement in neonatal body temperature during transportation to hospital, which were the outside temperature, gestational age, birth weight and the initial body temperature taken on arrival at the scene.

#### Complications

Supplementary Table 3 shows the wide-ranging types of complications encountered by emergency ambulance clinicians. The most common complications for the birth parent reported in the literature were PPH, abdominal pain, APH and breech presentations.

Premature birth, death/stillbirth, low birth weight, and hypothermia were most common for the neonate (Supplementary Table 3). Neonatal death and maternal cardiac arrest are likely to be over-represented in the literature in relation to the actual number of incidents which occur, as they are often reported as a single case study, which highlights the rarity and impact on clinicians.

#### Emergency ambulance clinician interventions

Many studies did not specifically report on interventions provided to patients. Supplementary Table 4 describes the management of OOHBs by emergency ambulance clinicians reported in the literature. Transportation to hospital and ‘vital signs’ were not included as a ‘management or intervention’ within the Table, as this would be relevant for most OOHBs.

### Birth parent, significant partner, and emergency ambulance clinician experiences with OOHBs

#### Birth parent & partner experiences

If a birth occurred in an unplanned location, birth parents (inclusive of significant partners) had often contacted the hospital within a few hours of the birth and experienced difficulty in determining the ‘right’ time to attend hospital.(6,30,68,70,71) Midwives triaging the birth parent frequently advised parents to stay at home until the labour was more advanced and were viewed as ‘gate keepers’ for admission, in some cases not sending an ambulance as requested.(69–71) The most common reasons surmised for an OOHB occurring were that for some birth parents the labour was not as painful as expected, or simply occurred precipitously, sometimes enroute to hospital.(6,56,68–72) Some parents deliberately delayed going to hospital to minimise interventions and have a more ‘natural’ birth, while others were organising suitable childcare for older siblings.(69,71)

Once it was apparent birth would occur outside the hospital, birth parents and partners often reported initial feelings of anxiety which were overtaken with focussing on the birth, and ultimately a feeling of achievement or empowerment following the birth.(6,68–71) Challenges such as no pain relief, blood loss, taxing environments (e.g.: car, ambulance), possible complications, and risk of hypothermia were often discussed.(6,56,68–72) Feelings of shame or guilt at not getting to the hospital before birth was reported by a few parents, or reliving what could have gone wrong, (6,68–71) while some were reconsidering future birth locations as they found the unplanned OOHB a positive empowering experience.(69,71) Partners who had previously been present at a birth reported this exposure assisted them with the OOHB as they knew what to expect. However, some partners would have liked to be more educated regarding what to do in the case of an OOHB occurring.(68,70)

Being coached through the birth via telephone was reassuring for many,(68,69,72) while one case was alarmed to be put on silence mode by a midwife during an en-caul birth.(6) Poor telephone networks affected some birth parent’s ability to contact the hospital or emergency ambulance when necessary.(69,72) Arrival of the ambulance was regarded as a relief and partners could shift their focus onto the birth parent. However, concerns were expressed by a number of birth parents about the apparent lack of training and expertise of ambulance clinicians, and in some cases the lack of respect, privacy, consideration of the birth plan, and consensual care which had implications for patient safety.(6,68,69,73) The emergency ambulance clinician’s communication skills and ability to place the birth parent at ease determined whether clinicians were perceived positively or negatively and could influence the tone of the OOHB experience.(6,68,70,73) Birth parents who perceived emergency ambulance clinician’s as unconfident interpreted the decision to provide rapid transportion to hospital as further evidence of low confidence, rather than being confident to allowing the birth to occur on scene.(8,73) Birth parents and partners reported being more relaxed if a midwife was present, and some expected a midwife to accompany emergency ambulance clinicians to a birth.(68,69)

Rural birth parents reported disappointment at centralisation of maternity services to regional areas requiring considerable travel to access. Cases have been reported where a birth parent was driving to hospital but had to stop due to advanced labour, and the ambulance had difficulty locating the patient due to unclear mapping.(69,72) Other birth parents described discomfort while being transported via ambulance while trying to safeguard their newborn.(70) Transfers could take longer than the birth parent had anticipated as well,(6) in some cases patients were diverted from one hospital to another.(70)

The hospital admission was usually regarded positively; however, a few birth parents described a lack of empathy from midwives who focussed more on the neonate or birth of the placenta. A need for debriefing was expressed, to understand what happened and allow the birth parent or partner to process the birth experience.(6,68–70)

#### Emergency ambulance clinician experiences

When emergency ambulance clinicians’ experiences were explored, many studies reported on the infrequency of OOHBs and a lack of training concerning obstetrics.(8,57,58,65) It was felt classroom learning did not translate well to real-life births where there is little clinical support, and often limited space and equipment and poor lighting and climate control.(65) Concerns were reported surrounding the limited opportunity for continuing education through clinical obstetrics placements.(8,58) Lack of exposure to births frequently led to low confidence in attending these cases.(8,57,58,65) A small number of services had midwives to assist for OOHB callouts while others could utilise telehealth for clinical support.(8,58)

OOHBs were often in challenging locations and the decision to stay on location or provide urgent transport to hospital is a critical clinical decision.(8,57,58) While it was recognised most OOHBs were uncomplicated, clinicians would ‘think through’ possible scenarios to best prepare themselves prior to arrival.(8,57,58) Uncomplicated births were a relief and viewed as a joyful occasion,(57,58) however complications were anxiety-provoking; some clinicians reported that due to lack of education/exposure they may be slower to recognise complications than is desirable, which could impact patient safety.(8,57) Prioritisation of care between the birth parent and neonate was difficult and some clinicians expressed concern handling a newborn.(57,65) Appropriate obstetrics and neonatal equipment were sometimes unavailable, for example neonatal thermometers and pulse oximetry capabilities.(57,58,65) Hill et al.(8) reported limited pain management options and fear of causing harm, especially to the neonate. Another study found if the neonate appeared well, no clinical observations may be undertaken, especially if the birth parent had complications.(65) Concerningly, optimal patient management may only occur if supported by clinical practice guidelines (guidelines may not support delayed cord cutting for example) or prompts provided in an electronic patient record.(8,65)

Additional challenges from clinicians’ perspectives included providing support to the family during birth or transport, long distances to definitive care, telecommunication difficulties, refusal to transport, demand for a female clinician to attend, language barriers, reliance on antenatal books for essential medical information, and lack of specialist clinical support.(8,57,58) Vagle et al.(58) suggested there may be a mismatch in the general public’s expectations of emergency ambulance clinicians’ role and skill level regarding births. Some clinicians desired a debriefing after attending an OOHB, especially if adverse outcomes occurred.(57,58) Handovers could be abrupt, and strained relations with midwives at the hospital were described.(57,58) There was a clear desire for high quality continuing education to enhance obstetric skills and knowledge.(8,57,58,65) Multiple authors (5,16,21,40,53,54,67,70,74) suggested there was a need to develop appropriate guidelines regarding OOHBs.

#### Barriers and challenges to optimal treatment provision for unplanned OOHBs and associated complications

Supplementary Table 5 includes the barriers and challenges identified within the 63 reviewed articles when attending OOHB’s. Identified barriers and challenges were varied, ranging from the desire for additional training, to revised OOHB guidelines, onto more practical issues such as minimal equipment appropriate for neonates being available on ambulances, and lack of role clarity when another clinician was also in attendance such as a midwife. The barriers and challenges have been broadly grouped into five categories: ‘education and training’, ‘communication and collaboration’, ‘environment’, ‘technology and aids’, and ‘other’.

## Discussion

This scoping review examined peer-reviewed research concerning imminent or OOHBs involving emergency ambulance clinician care. Emergency ambulance service data specifically analysing unplanned OOHBs, incidence, interventions, complications, and outcomes were uncommon within the literature. When reported, the incidence of OOHBs has remained low over time, at <1% of ambulance caseload.(3,16–20) However, recent international literature indicates an upwards trend of unplanned OOHBs, also influenced by the SARS-CoV-2 pandemic.(17,75) Townsend et al.(76) describes recent changes to pregnant patients’ engagement with healthcare systems worldwide, with reduced antenatal visits and an increase in virtual/remote care and unscheduled hospitalisations. Maternity unit closures in rural regions have further complicated access to maternity care, increasing OOHBs.(77) As these factors are influencing unplanned OOHBs, this creates an urgency in improving current emergency ambulance service training, education, and clinician confidence in this area.

For every OOHB there are ∼10 additional patients transported to hospital during labour.(2,3,16,19,50) Emergency ambulance clinicians scope of practice differs from midwives and often does not include skills such as foetal heart auscultation or confirming effacement, which determines whether the patient is in active second stage of labour.(3,22,42,50) Therefore, rapid history-taking and patient observation help inform the decision to provide urgent transport or stay on scene.(8) However, some studies did report on the ability to confirm dilation,(43,63,66) indicating differing roles and scope of practice worldwide. Whilst a majority of cases are precipitous and uncomplicated, as many as 28% of OOHBS may be premature(3) and 86% of neonates may experience hypothermia(40), although most studies reported lower incidences of these conditions. The most common maternal complication reported was PPH (Supplementary Table 3). PPH was often undefined in terms of the volume of blood loss,(16,20,65,21,22,41,46,48,52,59,60) while hypothermia definitions varied from <36.5 degrees to <36.0 degrees, creating some heterogeneity within results. Independent predictive risk factors for neonatal morbidity and mortality following an OOHB include multiparity, maternal pathology, prematurity and neonatal hypothermia.(5) OOH management of PPH and hypothermia may not be as effective as in-hospital care, which increases the risk of adverse outcomes for birth parent and neonate.(29,32,33,37–41,45–47,65,67) In some countries midwives accompany ambulance personnel for OOHBs,(6,60) and in France mobile intensive care units (MICU) respond to these requests comprising of a driver, nurse and senior emergency doctor.(29). This contrasts to the Australian and American systems where paramedics, or in some regions, volunteer ambulance officers, attend OOH emergencies (including OOHBs),(78,79) although, it is not uncommon for a medical doctor to attend during aeromedical transfers in Australia.(28) Unsurprisingly, OOHBs and associated complications have improved outcomes if a skilled clinician is present.(35,80,81) Given the centralisation of many maternity services, this poses considerable problems for rural and remote regions where there may be large distances to definitive care.(21,57,82–84) Aeromedical retrieval is sometimes an option but is subject to availability and weather conditions, so may not always be viable.(28,63,66)

Limited studies report on OOHBs from the patient, bystander or clinician perspective. There is a significant gap in the research pertaining to birth parent and partner’s experience of adverse outcomes (acknowledging sensitivity/privacy considerations); nevertheless, understanding the more complex scenarios is important as this may provide compelling evidence for service improvement, guideline revision, and training. Some emergency ambulance clinician’s describe serious adverse outcomes when attending OOHBs and how harrowing this is,(8,58) with little discussion provided in the literature regarding organisational support following a difficult case.(25,27) It is unclear how often organisational support services are accessed, whether this is actively encouraged by organisations, and whether this benefits the clinician if the services are utilised. Occupational stress and trauma experienced by these clinicians is worthy of further research to assess the impact of trauma on individuals and organisations, and how to reduce the possibility of long-term stress disorders and ill health in this valuable resource.

Significant barriers remain for treatment and management of OOHB patients. Many of these challenges are inherent to any OOH situation, such as difficult extrications, weather, language difficulties or bystanders complicating scenes. Specific to OOHBs, collaboration between maternity services, receiving hospitals, and ambulance services appears to be problematic in certain countries or regions,(36,53,55,57,58,74) in addition to lack of clarity regarding roles and responsibilities when a midwife is also in attendance.(60,65) Previously, interfacility training has proven beneficial for both students and experienced clinicians in understanding roles, improving communication, and facilitating learning transfer between midwives and paramedics concerning OOHBs.(85–87) The rarity of these events has raised concerns regarding emergency ambulance clinician knowledge and skill decay, with multiple authors recommending improvements in ongoing training and education provision to clinicians to better retain skills currency.(Supplementary Table 5)

## Strengths and limitations

This review provides a comprehensive overview of imminent births and unplanned OOHBs attended by emergency ambulance clinicians in high income countries. This also includes planned home births or freebirths where emergency ambulance assistance is required. A rigorous methodology framework was utilised including the JBI and PRISMA-ScR guidelines.(9–12) This review did not consider risk of bias, rigour, or quality of included studies, which is not uncommon for scoping reviews.(10) However, most research in this space was identified as using retrospective data of patient records or qualitative research methodologies, which is considered a lower level of evidence.(88) Five databases were reviewed by one author using the search strategy described in Figure 1, and located articles were corroborated for inclusion by a second author. However, any consideration of limitations must include the possibility of missed articles. Additionally, only articles published in English were included, excluding data from non-English publications. Nevertheless, this review provides a comprehensive review of available literature pertaining to OOH involvement in OOHBs or where there is potential for an OOHB in paramedic care.

## Conclusion

Reviewing 63 articles concerning emergency ambulance clinician involvement in OOHBs, it is apparent these situations remain a low-frequency, high-risk event. The majority of articles were drawn from the USA, Australia, and United Kingdom, and included over 40,596 unplanned OOHBs. The literature found 51 factors which increased the risk of an OOHB, and 26 factors which increase the incidence of neonatal morbidity and mortality. Emergency ambulance clinicians encountered 99 peripartum complications with 32 interventions utilised for OOHBs. Usually birth parent, partner and clinician experienced initial anxiety before the birth, however this turned to focussing on the birth, followed by relief and joy if a healthy neonate was born. Many challenges exist to optimal management of OOHBs, broadly encompassing communication, collaboration with other healthcare professionals (such as midwives or emergency department clinicians), environmental and technological issues. It is reported that many emergency ambulance clinicians would benefit from ongoing training and education pertaining to these rare events to improve patient outcomes.

## Supporting information

PRISMA-ScR check list

## Author Contributions

Concept and design of the research: MH, BM, LH, SH, AM and BF.

Screened articles for inclusion criteria: MH, AM.

Data extraction and consistency of included data: MH, BM. Draft article: MH.

Revisions: AM, SH, BM, BF and LH.

Content expert: BF.

## Funding

This research was undertaken with assistance from a PhD scholarship awarded as part of Dr Luke Hopper’s ECU Vice Chancellor’s Research Fellow Scholarship for 2020. There are no grant/award numbers associated with this.

## Competing interests

None declared.

## Patient consent

Not required

## Abbreviations

BBA: Born before arrival (or birth before arrival)

JBI: Joanna Briggs Institute

OOH: Out-of-hospital

OOHB: Out-of-hospital birth

PPH: Postpartum haemorrhage

PRISMA-ScR: Preferred Reporting Items for Systematic Reviews and Meta-Analysis extension for Scoping Reviews

UK: United Kingdom

USA: United State of America

## Data Availability

All data produced in the present work are contained in the manuscript

---

**Supplementary Table 1:**
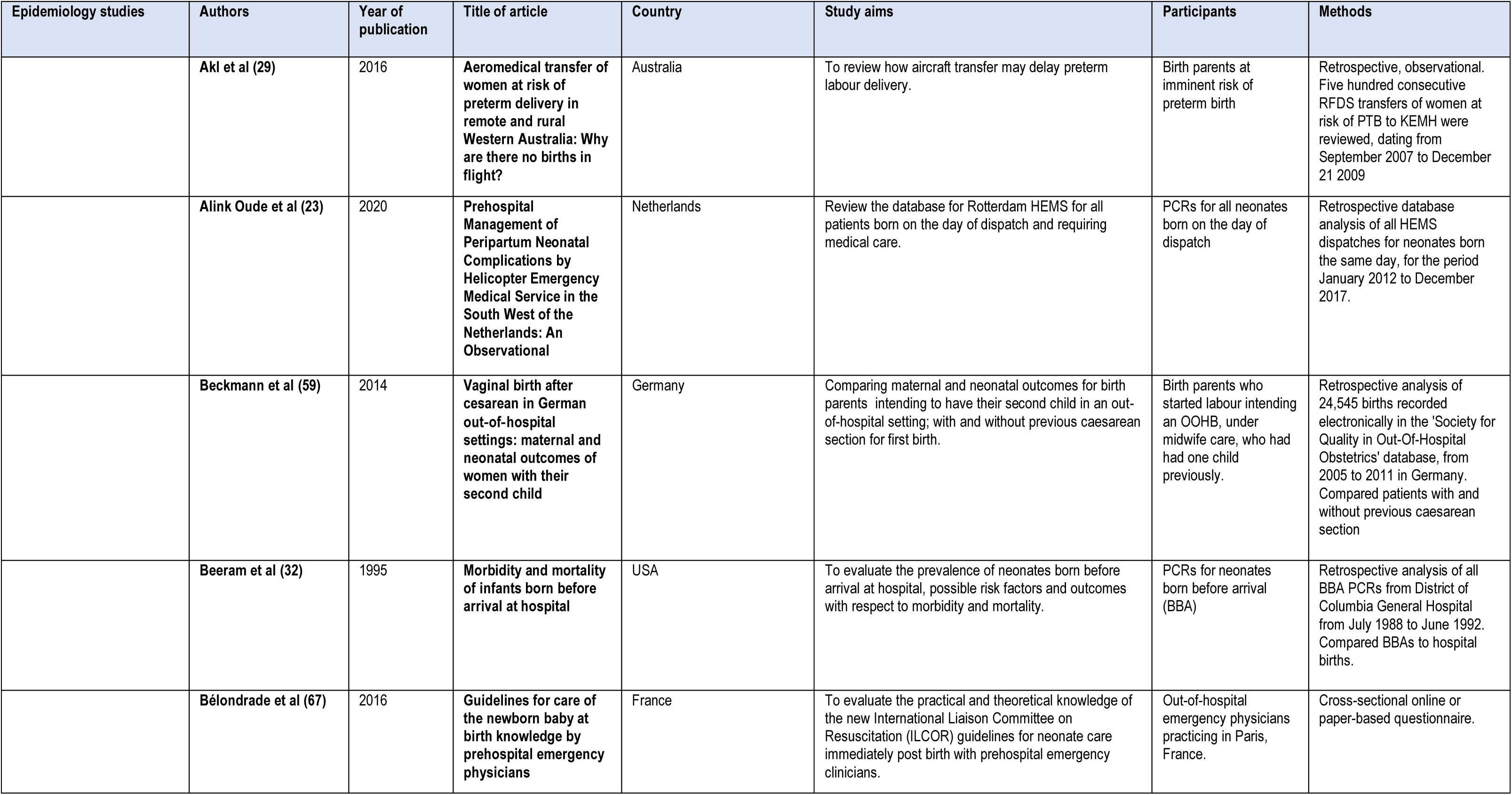
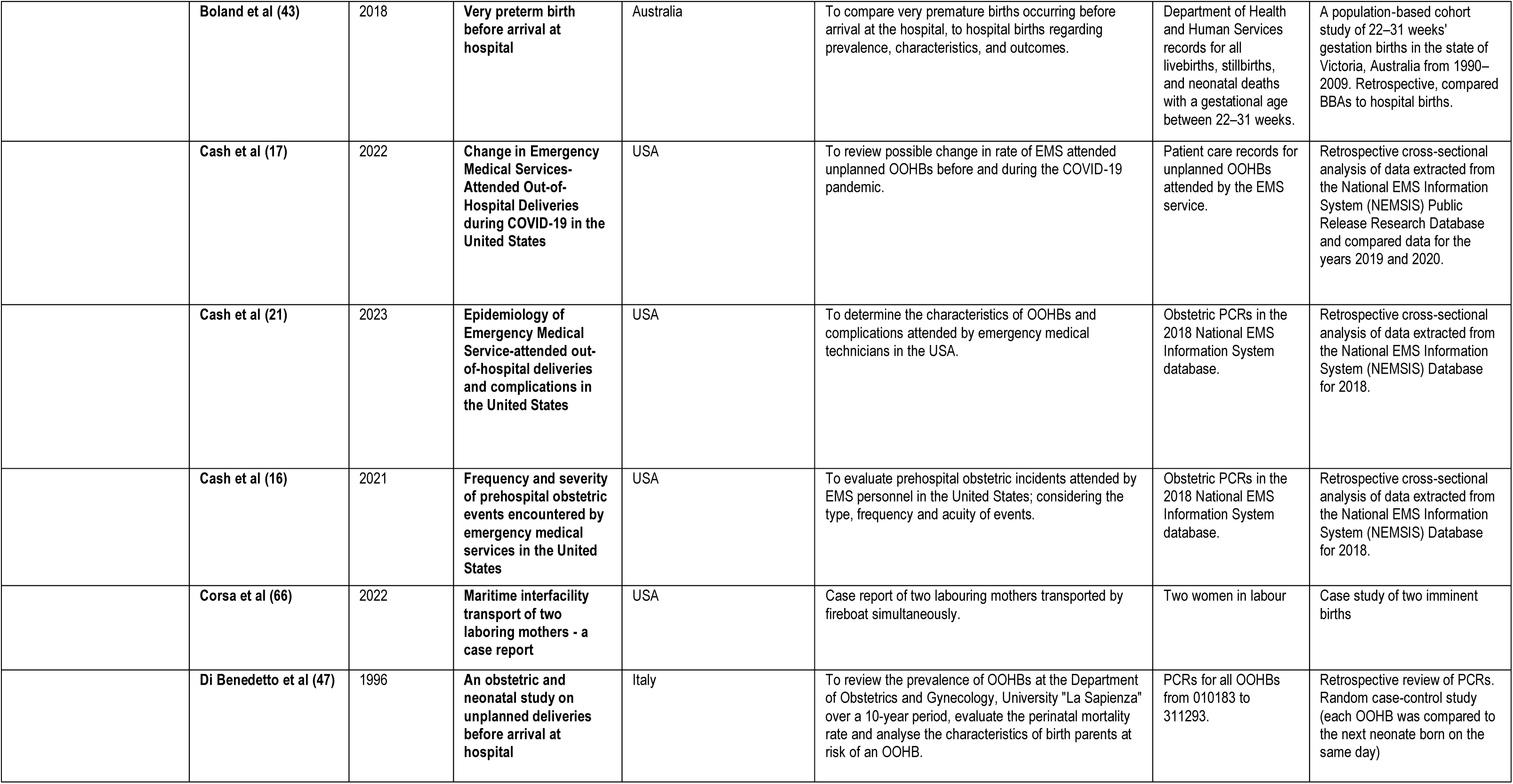
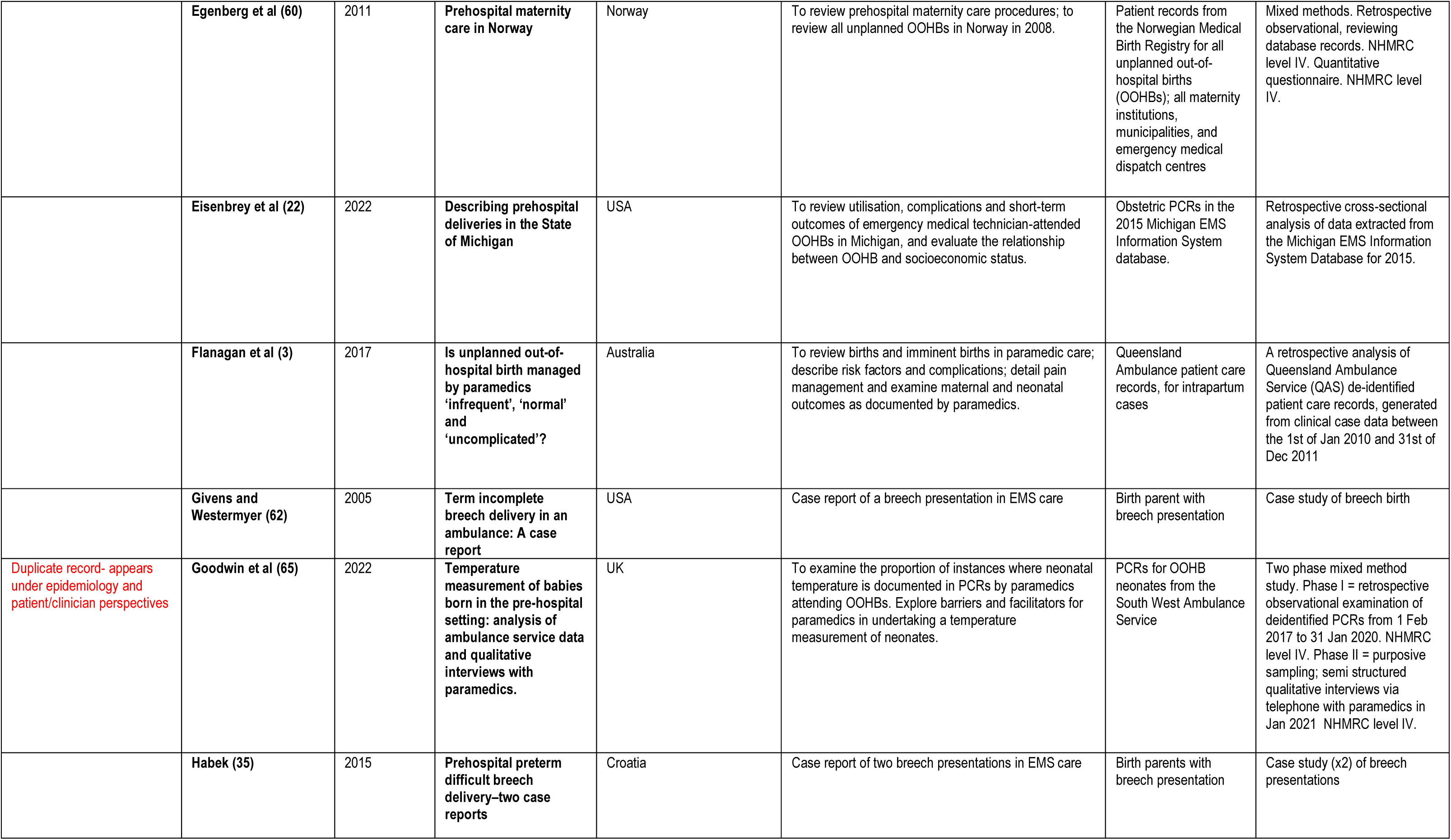
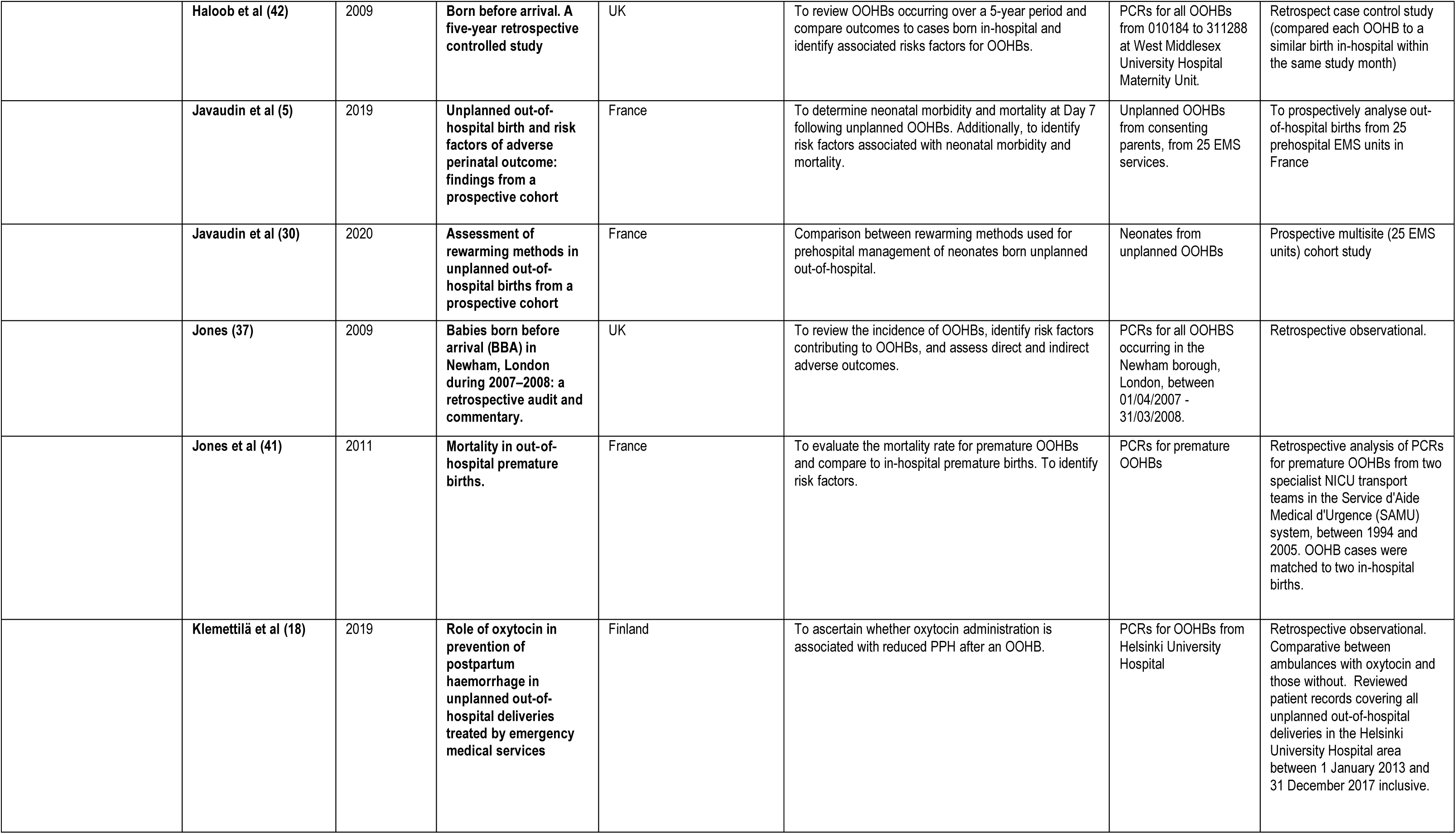
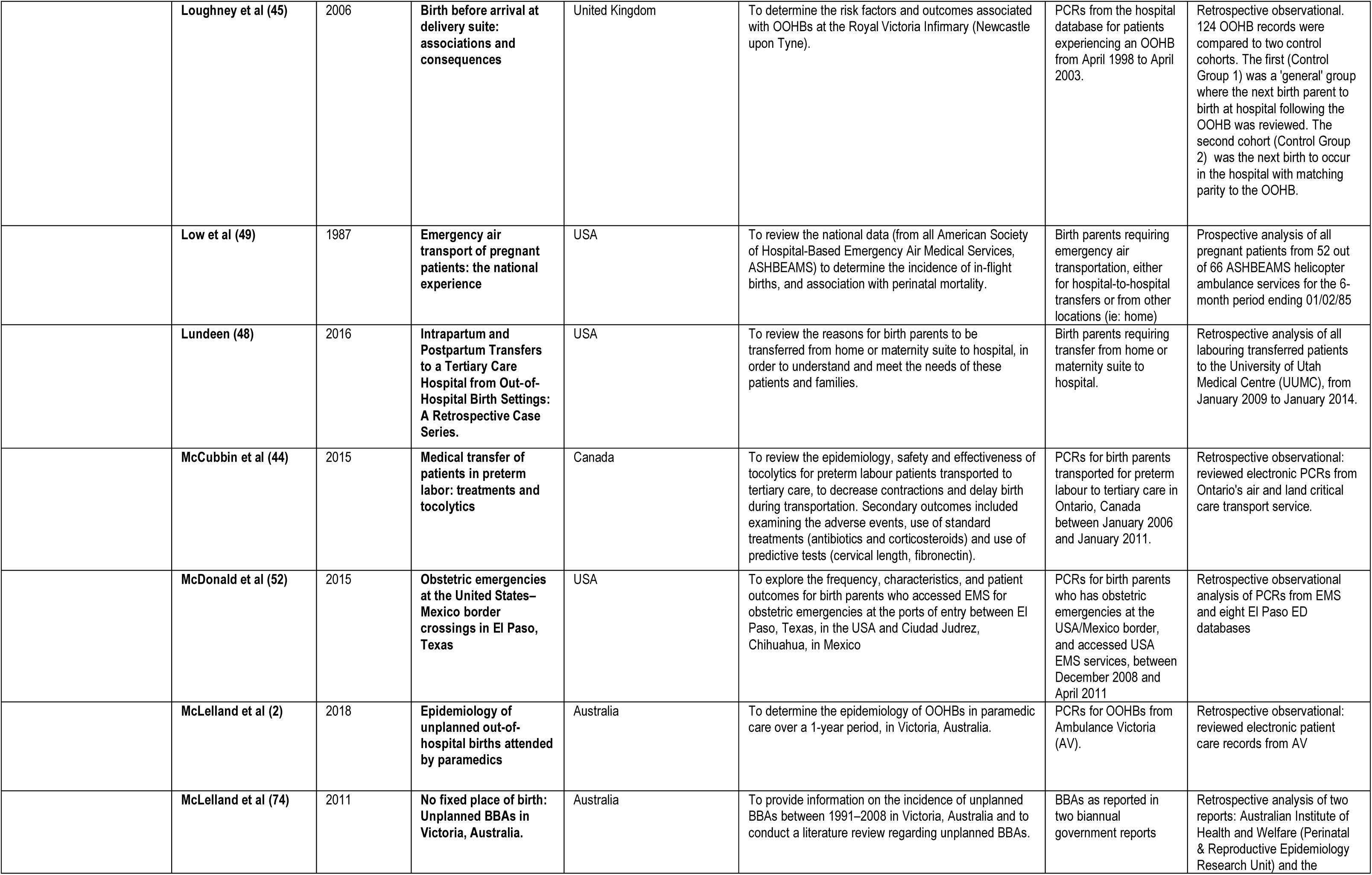
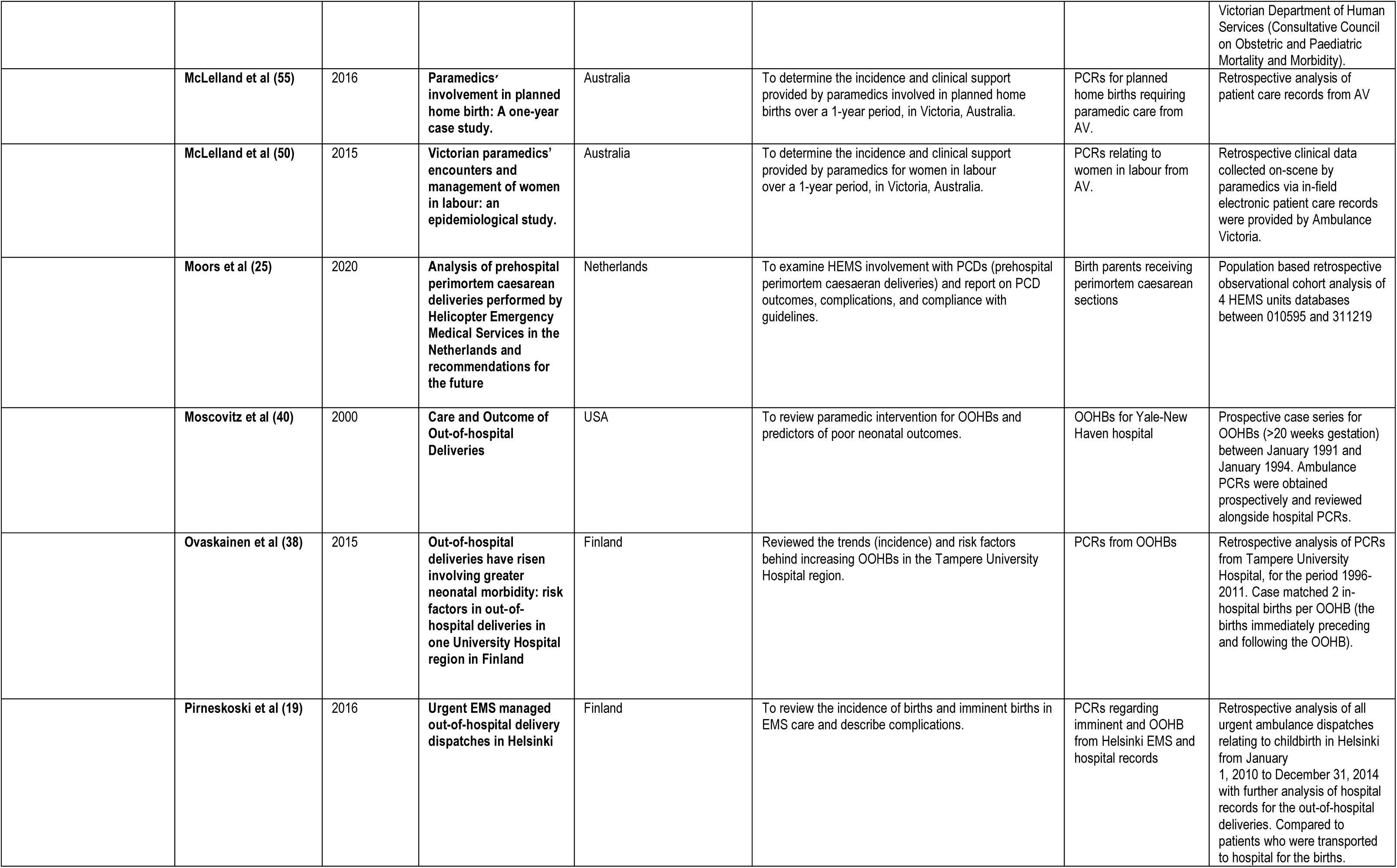
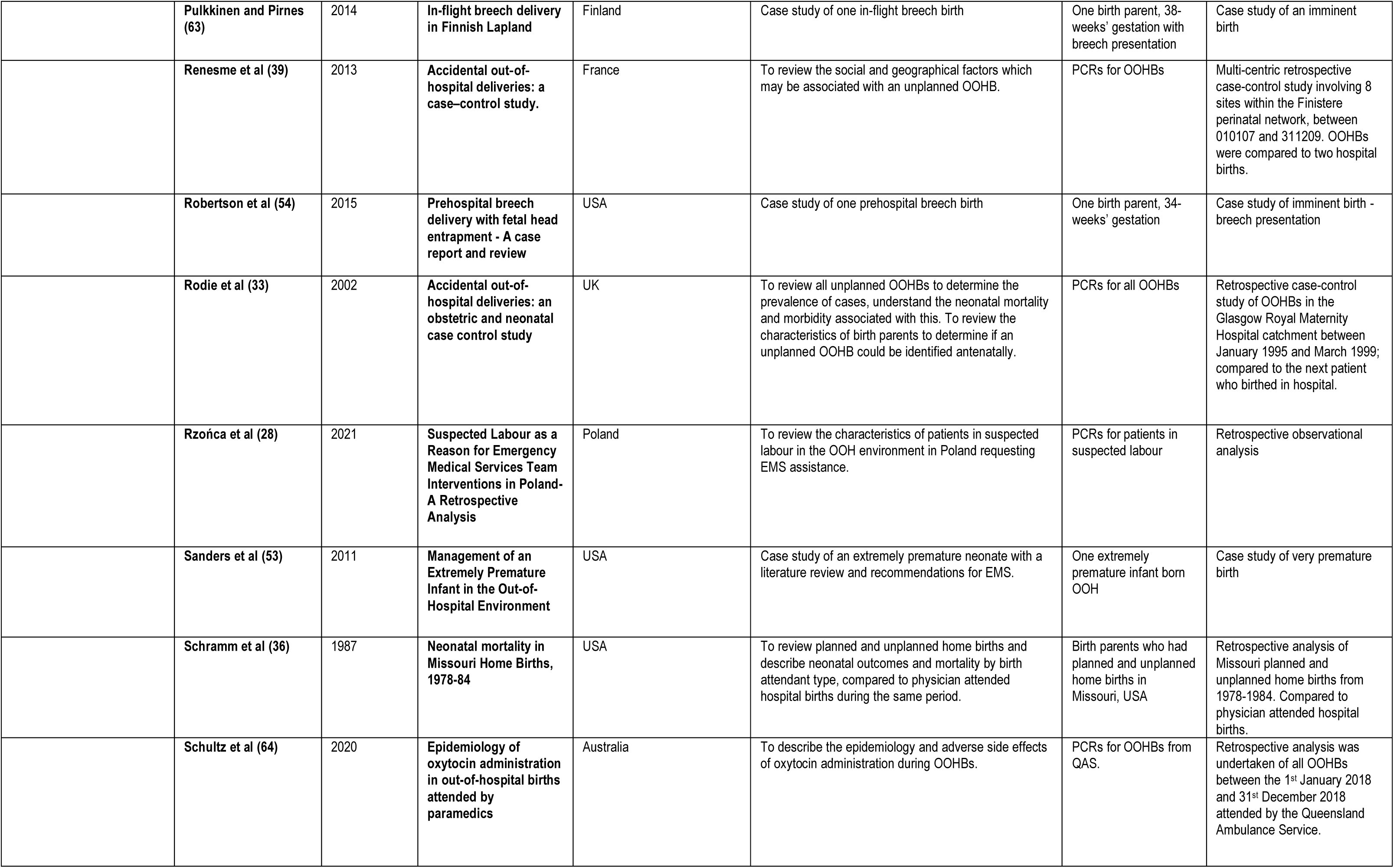
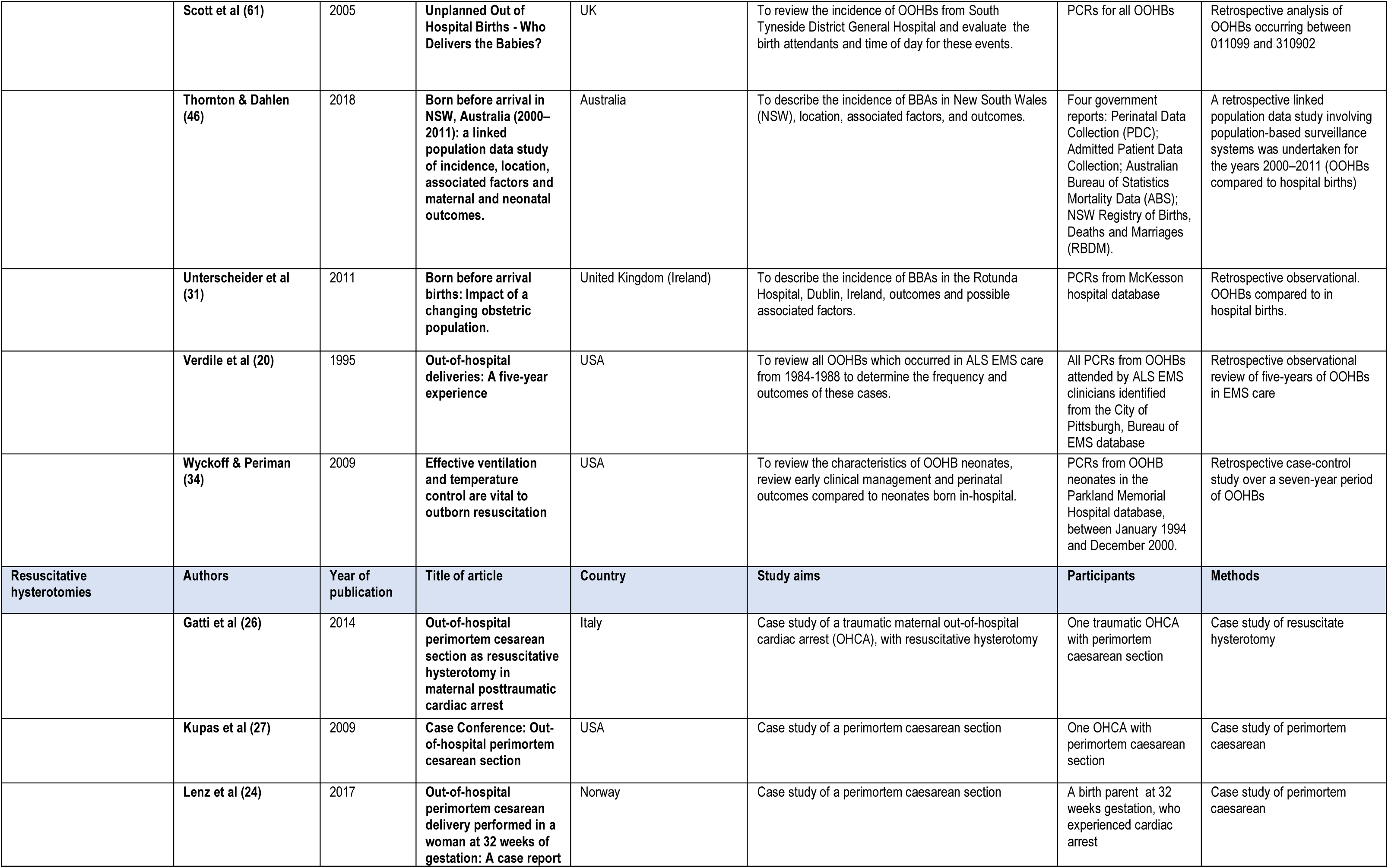
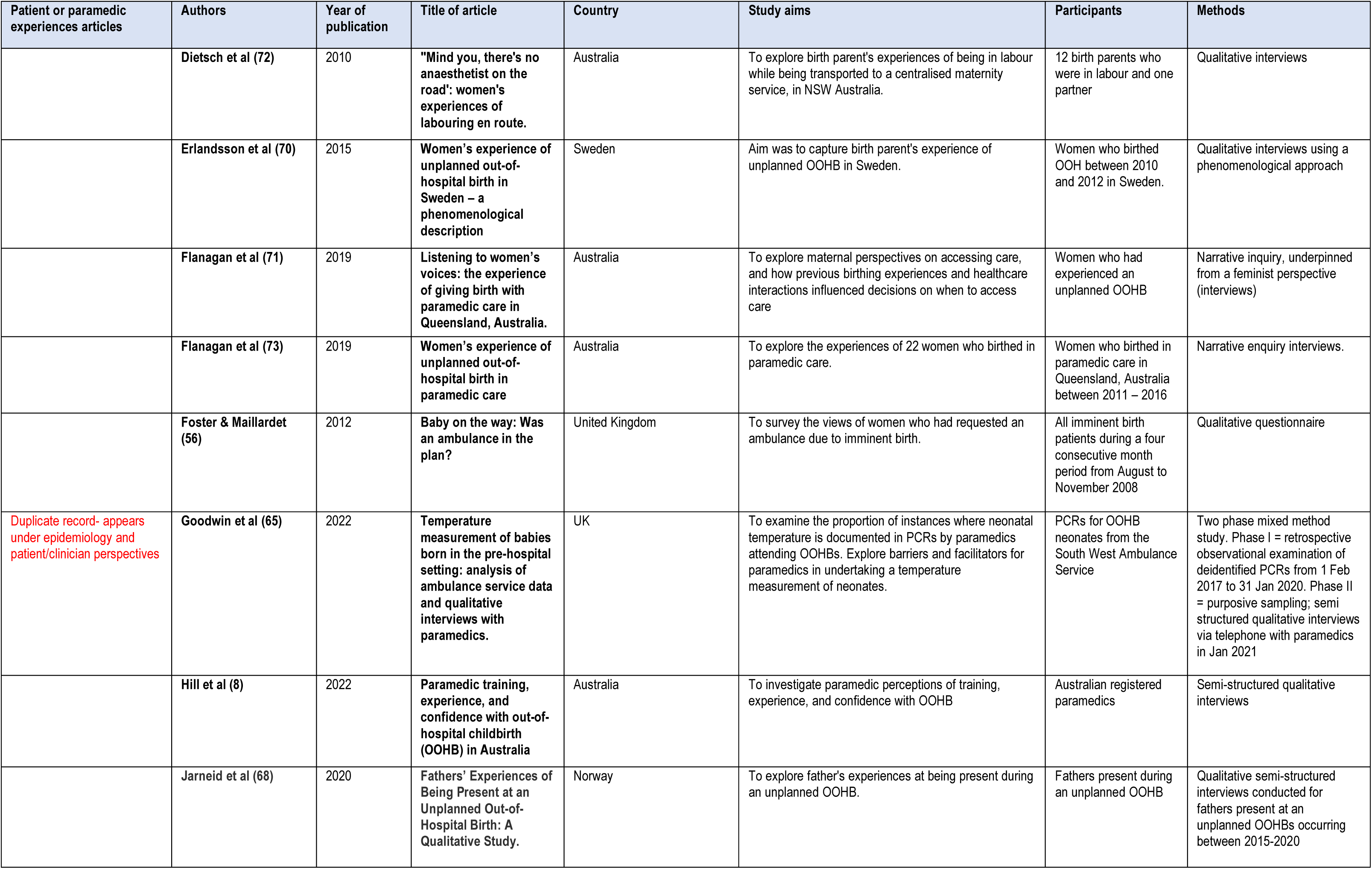
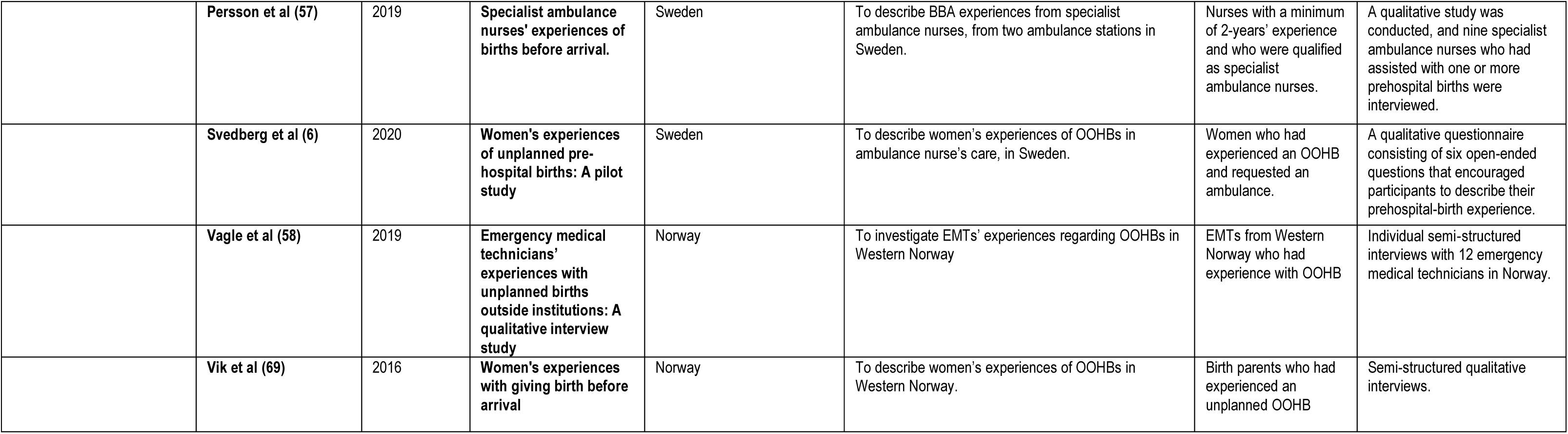
Included articles for Out-of-hospital births and experiences of emergency ambulance clinicians and birthing parents: a scoping review of the literature.

| Epidemiology studies | Authors | Year of publication | Title of article | Country | Study aims | Participants | Methods |
| --- | --- | --- | --- | --- | --- | --- | --- |
|  | Akl et al (29) | 2016 | Aeromedical transfer of women at risk of preterm delivery in remote and rural Western Australia: Why are there no births in flight? | Australia | To review how aircraft transfer may delay preterm labour delivery. | Birth parents at imminent risk of preterm birth | Retrospective, observational. Five hundred consecutive RFDS transfers of women at risk of PTB to KEMH were reviewed, dating from September 2007 to December 21 2009 |
|  | Alink Oude et al (23) | 2020 | Prehospital Management of Peripartum Neonatal Complications by Helicopter Emergency Medical Service in the South West of the Netherlands: An Observational | Netherlands | Review the database for Rotterdam HEMS for all patients born on the day of dispatch and requiring medical care. | PCRs for all neonates born on the day of dispatch | Retrospective database analysis of all HEMS dispatches for neonates born the same day, for the period January 2012 to December 2017. |
|  | Beckmann et al (59) | 2014 | Vaginal birth after cesarean in German out-of-hospital settings: maternal and neonatal outcomes of women with their second child | Germany | Comparing maternal and neonatal outcomes for birth parents intending to have their second child in an out-of-hospital setting; with and without previous caesarean section for first birth. | Birth parents who started labour intending an OOHb, under midwife care, who had had one child previously. | Retrospective analysis of 24,545 births recorded electronically in the 'Society for Quality in Out-Of-Hospital Obstetrics' database, from 2005 to 2011 in Germany. Compared patients with and without previous caesarean section |
|  | Beeram et al (32) | 1995 | Morbidity and mortality of infants born before arrival at hospital | USA | To evaluate the prevalence of neonates born before arrival at hospital, possible risk factors and outcomes with respect to morbidity and mortality. | PCRs for neonates born before arrival (BBA) | Retrospective analysis of all BBA PCRs from District of Columbia General Hospital from July 1988 to June 1992. Compared BBAs to hospital births. |
|  | Bélondrade et al (67) | 2016 | Guidelines for care of the newborn baby at birth knowledge by prehospital emergency physicians | France | To evaluate the practical and theoretical knowledge of the new International Liaison Committee on Resuscitation (ILCOR) guidelines for neonate care immediately post birth with prehospital emergency clinicians. | Out-of-hospital emergency physicians practicing in Paris, France. | Cross-sectional online or paper-based questionnaire. |
|  | **Boland et al (43)** | 2018 | **Very preterm birth before arrival at hospital** | Australia | To compare very premature births occurring before arrival at the hospital, to hospital births regarding prevalence, characteristics, and outcomes. | Department of Health and Human Services records for all livebirths, stillbirths, and neonatal deaths with a gestational age between 22–31 weeks. | A population-based cohort study of 22–31 weeks' gestation births in the state of Victoria, Australia from 1990–2009. Retrospective, compared BBAs to hospital births. |
|  | **Cash et al (17)** | 2022 | **Change in Emergency Medical Services-Attended Out-of-Hospital Deliveries during COVID-19 in the United States** | USA | To review possible change in rate of EMS attended unplanned OOHs before and during the COVID-19 pandemic. | Patient care records for unplanned OOHs attended by the EMS service. | Retrospective cross-sectional analysis of data extracted from the National EMS Information System (NEMSIS) Public Release Research Database and compared data for the years 2019 and 2020. |
|  | **Cash et al (21)** | 2023 | **Epidemiology of Emergency Medical Service-attended out-of-hospital deliveries and complications in the United States** | USA | To determine the characteristics of OOHs and complications attended by emergency medical technicians in the USA. | Obstetric PCRs in the 2018 National EMS Information System database. | Retrospective cross-sectional analysis of data extracted from the National EMS Information System (NEMSIS) Database for 2018. |
|  | **Cash et al (16)** | 2021 | **Frequency and severity of prehospital obstetric events encountered by emergency medical services in the United States** | USA | To evaluate prehospital obstetric incidents attended by EMS personnel in the United States; considering the type, frequency and acuity of events. | Obstetric PCRs in the 2018 National EMS Information System database. | Retrospective cross-sectional analysis of data extracted from the National EMS Information System (NEMSIS) Database for 2018. |
|  | **Corsa et al (66)** | 2022 | **Maritime interfacility transport of two laboring mothers - a case report** | USA | Case report of two labouring mothers transported by fireboat simultaneously. | Two women in labour | Case study of two imminent births |
|  | **Di Benedetto et al (47)** | 1996 | **An obstetric and neonatal study on unplanned deliveries before arrival at hospital** | Italy | To review the prevalence of OOHs at the Department of Obstetrics and Gynecology, University "La Sapienza" over a 10-year period, evaluate the perinatal mortality rate and analyse the characteristics of birth parents at risk of an OOH. | PCRs for all OOHs from 010183 to 311293. | Retrospective review of PCRs. Random case-control study (each OOH was compared to the next neonate born on the same day) |
|  | Egenberg et al (60) | 2011 | Prehospital maternity care in Norway | Norway | To review prehospital maternity care procedures; to review all unplanned OOHs in Norway in 2008. | Patient records from the Norwegian Medical Birth Registry for all unplanned out-of-hospital births (OOHBs); all maternity institutions, municipalities, and emergency medical dispatch centres | Mixed methods. Retrospective observational, reviewing database records. NHMRC level IV. Quantitative questionnaire. NHMRC level IV. |
|  | Eisenbrey et al (22) | 2022 | Describing prehospital deliveries in the State of Michigan | USA | To review utilisation, complications and short-term outcomes of emergency medical technician-attended OOHs in Michigan, and evaluate the relationship between OOHb and socioeconomic status. | Obstetric PCRs in the 2015 Michigan EMS Information System database. | Retrospective cross-sectional analysis of data extracted from the Michigan EMS Information System Database for 2015. |
|  | Flanagan et al (3) | 2017 | Is unplanned out-of-hospital birth managed by paramedics 'infrequent', 'normal' and 'uncomplicated'? | Australia | To review births and imminent births in paramedic care; describe risk factors and complications; detail pain management and examine maternal and neonatal outcomes as documented by paramedics. | Queensland Ambulance patient care records, for intrapartum cases | A retrospective analysis of Queensland Ambulance Service (QAS) de-identified patient care records, generated from clinical case data between the 1st of Jan 2010 and 31st of Dec 2011 |
|  | Givens and Westermeyer (62) | 2005 | Term incomplete breech delivery in an ambulance: A case report | USA | Case report of a breech presentation in EMS care | Birth parent with breech presentation | Case study of breech birth |
| Duplicate record- appears under epidemiology and patient/clinician perspectives | Goodwin et al (65) | 2022 | Temperature measurement of babies born in the pre-hospital setting: analysis of ambulance service data and qualitative interviews with paramedics. | UK | To examine the proportion of instances where neonatal temperature is documented in PCRs by paramedics attending OOHs. Explore barriers and facilitators for paramedics in undertaking a temperature measurement of neonates. | PCRs for OOHb neonates from the South West Ambulance Service | Two phase mixed method study. Phase I = retrospective observational examination of deidentified PCRs from 1 Feb 2017 to 31 Jan 2020. NHMRC level IV. Phase II = purposive sampling; semi structured qualitative interviews via telephone with paramedics in Jan 2021 NHMRC level IV. |
|  | Habek (35) | 2015 | Prehospital preterm difficult breech delivery—two case reports | Croatia | Case report of two breech presentations in EMS care | Birth parents with breech presentation | Case study (x2) of breech presentations |
|  | **Haloob et al (42)** | 2009 | **Born before arrival. A five-year retrospective controlled study** | UK | To review OOHBs occurring over a 5-year period and compare outcomes to cases born in-hospital and identify associated risks factors for OOHBs. | PCRs for all OOHBs from 010184 to 311288 at West Middlesex University Hospital Maternity Unit. | Retrospect case control study (compared each OOHb to a similar birth in-hospital within the same study month) |
|  | **Javaudin et al (5)** | 2019 | **Unplanned out-of-hospital birth and risk factors of adverse perinatal outcome: findings from a prospective cohort** | France | To determine neonatal morbidity and mortality at Day 7 following unplanned OOHBs. Additionally, to identify risk factors associated with neonatal morbidity and mortality. | Unplanned OOHBs from consenting parents, from 25 EMS services. | To prospectively analyse out-of-hospital births from 25 prehospital EMS units in France |
|  | **Javaudin et al (30)** | 2020 | **Assessment of rewarming methods in unplanned out-of-hospital births from a prospective cohort** | France | Comparison between rewarming methods used for prehospital management of neonates born unplanned out-of-hospital. | Neonates from unplanned OOHBs | Prospective multisite (25 EMS units) cohort study |
|  | **Jones (37)** | 2009 | **Babies born before arrival (BBA) in Newham, London during 2007–2008: a retrospective audit and commentary.** | UK | To review the incidence of OOHBs, identify risk factors contributing to OOHBs, and assess direct and indirect adverse outcomes. | PCRs for all OOHBS occurring in the Newham borough, London, between 01/04/2007 - 31/03/2008. | Retrospective observational. |
|  | **Jones et al (41)** | 2011 | **Mortality in out-of-hospital premature births.** | France | To evaluate the mortality rate for premature OOHBs and compare to in-hospital premature births. To identify risk factors. | PCRs for premature OOHBs | Retrospective analysis of PCRs for premature OOHBs from two specialist NICU transport teams in the Service d'Aide Medical d'Urgence (SAMU) system, between 1994 and 2005. OOHb cases were matched to two in-hospital births. |
|  | **Klemettilä et al (18)** | 2019 | **Role of oxytocin in prevention of postpartum haemorrhage in unplanned out-of-hospital deliveries treated by emergency medical services** | Finland | To ascertain whether oxytocin administration is associated with reduced PPH after an OOHb. | PCRs for OOHBs from Helsinki University Hospital | Retrospective observational. Comparative between ambulances with oxytocin and those without. Reviewed patient records covering all unplanned out-of-hospital deliveries in the Helsinki University Hospital area between 1 January 2013 and 31 December 2017 inclusive. |
|  | **Loughney et al (45)** | 2006 | **Birth before arrival at delivery suite: associations and consequences** | United Kingdom | To determine the risk factors and outcomes associated with OOHBs at the Royal Victoria Infirmary (Newcastle upon Tyne). | PCRs from the hospital database for patients experiencing an OOHb from April 1998 to April 2003. | Retrospective observational. 124 OOHb records were compared to two control cohorts. The first (Control Group 1) was a 'general' group where the next birth parent to birth at hospital following the OOHb was reviewed. The second cohort (Control Group 2) was the next birth to occur in the hospital with matching parity to the OOHb. |
|  | **Low et al (49)** | 1987 | **Emergency air transport of pregnant patients: the national experience** | USA | To review the national data (from all American Society of Hospital-Based Emergency Air Medical Services, ASHBEAMS) to determine the incidence of in-flight births, and association with perinatal mortality. | Birth parents requiring emergency air transportation, either for hospital-to-hospital transfers or from other locations (ie: home) | Prospective analysis of all pregnant patients from 52 out of 66 ASHBEAMS helicopter ambulance services for the 6-month period ending 01/02/85 |
|  | **Lundeen (48)** | 2016 | **Intrapartum and Postpartum Transfers to a Tertiary Care Hospital from Out-of-Hospital Birth Settings: A Retrospective Case Series.** | USA | To review the reasons for birth parents to be transferred from home or maternity suite to hospital, in order to understand and meet the needs of these patients and families. | Birth parents requiring transfer from home or maternity suite to hospital. | Retrospective analysis of all labouring transferred patients to the University of Utah Medical Centre (UUMC), from January 2009 to January 2014. |
|  | **McCubbin et al (44)** | 2015 | **Medical transfer of patients in preterm labor: treatments and tocolytics** | Canada | To review the epidemiology, safety and effectiveness of tocolytics for preterm labour patients transported to tertiary care, to decrease contractions and delay birth during transportation. Secondary outcomes included examining the adverse events, use of standard treatments (antibiotics and corticosteroids) and use of predictive tests (cervical length, fibronectin). | PCRs for birth parents transported for preterm labour to tertiary care in Ontario, Canada between January 2006 and January 2011. | Retrospective observational: reviewed electronic PCRs from Ontario's air and land critical care transport service. |
|  | **McDonald et al (52)** | 2015 | **Obstetric emergencies at the United States–Mexico border crossings in El Paso, Texas** | USA | To explore the frequency, characteristics, and patient outcomes for birth parents who accessed EMS for obstetric emergencies at the ports of entry between El Paso, Texas, in the USA and Ciudad Juarez, Chihuahua, in Mexico | PCRs for birth parents who has obstetric emergencies at the USA/Mexico border, and accessed USA EMS services, between December 2008 and April 2011 | Retrospective observational analysis of PCRs from EMS and eight El Paso ED databases |
|  | **McLelland et al (2)** | 2018 | **Epidemiology of unplanned out-of-hospital births attended by paramedics** | Australia | To determine the epidemiology of OOHBs in paramedic care over a 1-year period, in Victoria, Australia. | PCRs for OOHBs from Ambulance Victoria (AV). | Retrospective observational: reviewed electronic patient care records from AV |
|  | **McLelland et al (74)** | 2011 | **No fixed place of birth: Unplanned BBAs in Victoria, Australia.** | Australia | To provide information on the incidence of unplanned BBAs between 1991–2008 in Victoria, Australia and to conduct a literature review regarding unplanned BBAs. | BBAs as reported in two biannual government reports | Retrospective analysis of two reports: Australian Institute of Health and Welfare (Perinatal & Reproductive Epidemiology Research Unit) and the |
|  |  |  |  |  |  |  | Victorian Department of Human Services (Consultative Council on Obstetric and Paediatric Mortality and Morbidity). |
|  | McLelland et al (55) | 2016 | Paramedics' involvement in planned home birth: A one-year case study. | Australia | To determine the incidence and clinical support provided by paramedics involved in planned home births over a 1-year period, in Victoria, Australia. | PCRs for planned home births requiring paramedic care from AV. | Retrospective analysis of patient care records from AV |
|  | McLelland et al (50) | 2015 | Victorian paramedics' encounters and management of women in labour: an epidemiological study. | Australia | To determine the incidence and clinical support provided by paramedics for women in labour over a 1-year period, in Victoria, Australia. | PCRs relating to women in labour from AV. | Retrospective clinical data collected on-scene by paramedics via in-field electronic patient care records were provided by Ambulance Victoria. |
|  | Moors et al (25) | 2020 | Analysis of prehospital perimortem caesarean deliveries performed by Helicopter Emergency Medical Services in the Netherlands and recommendations for the future | Netherlands | To examine HEMS involvement with PCDs (prehospital perimortem caesarean deliveries) and report on PCD outcomes, complications, and compliance with guidelines. | Birth parents receiving perimortem caesarean sections | Population based retrospective observational cohort analysis of 4 HEMS units databases between 010595 and 311219 |
|  | Moscovitz et al (40) | 2000 | Care and Outcome of Out-of-hospital Deliveries | USA | To review paramedic intervention for OOHBs and predictors of poor neonatal outcomes. | OOHBs for Yale-New Haven hospital | Prospective case series for OOHBs (>20 weeks gestation) between January 1991 and January 1994. Ambulance PCRs were obtained prospectively and reviewed alongside hospital PCRs. |
|  | Ovaskainen et al (38) | 2015 | Out-of-hospital deliveries have risen involving greater neonatal morbidity: risk factors in out-of-hospital deliveries in one University Hospital region in Finland | Finland | Reviewed the trends (incidence) and risk factors behind increasing OOHBs in the Tampere University Hospital region. | PCRs from OOHBs | Retrospective analysis of PCRs from Tampere University Hospital, for the period 1996-2011. Case matched 2 in-hospital births per OOHb (the births immediately preceding and following the OOHb). |
|  | Pirneskoski et al (19) | 2016 | Urgent EMS managed out-of-hospital delivery dispatches in Helsinki | Finland | To review the incidence of births and imminent births in EMS care and describe complications. | PCRs regarding imminent and OOHb from Helsinki EMS and hospital records | Retrospective analysis of all urgent ambulance dispatches relating to childbirth in Helsinki from January 1, 2010 to December 31, 2014 with further analysis of hospital records for the out-of-hospital deliveries. Compared to patients who were transported to hospital for the births. |
|  | **Pulkkinen and Pirnes (63)** | 2014 | **In-flight breech delivery in Finnish Lapland** | Finland | Case study of one in-flight breech birth | One birth parent, 38-weeks' gestation with breech presentation | Case study of an imminent birth |
|  | **Renesme et al (39)** | 2013 | **Accidental out-of-hospital deliveries: a case-control study.** | France | To review the social and geographical factors which may be associated with an unplanned OOHb. | PCRs for OOHbs | Multi-centric retrospective case-control study involving 8 sites within the Finistere perinatal network, between 010107 and 311209. OOHbs were compared to two hospital births. |
|  | **Robertson et al (54)** | 2015 | **Prehospital breech delivery with fetal head entrapment - A case report and review** | USA | Case study of one prehospital breech birth | One birth parent, 34-weeks' gestation | Case study of imminent birth - breech presentation |
|  | **Rodie et al (33)** | 2002 | **Accidental out-of-hospital deliveries: an obstetric and neonatal case control study** | UK | To review all unplanned OOHbs to determine the prevalence of cases, understand the neonatal mortality and morbidity associated with this. To review the characteristics of birth parents to determine if an unplanned OOHb could be identified antenatally. | PCRs for all OOHbs | Retrospective case-control study of OOHbs in the Glasgow Royal Maternity Hospital catchment between January 1995 and March 1999; compared to the next patient who birthed in hospital. |
|  | **Rzońca et al (28)** | 2021 | **Suspected Labour as a Reason for Emergency Medical Services Team Interventions in Poland- A Retrospective Analysis** | Poland | To review the characteristics of patients in suspected labour in the OOH environment in Poland requesting EMS assistance. | PCRs for patients in suspected labour | Retrospective observational analysis |
|  | **Sanders et al (53)** | 2011 | **Management of an Extremely Premature Infant in the Out-of-Hospital Environment** | USA | Case study of an extremely premature neonate with a literature review and recommendations for EMS. | One extremely premature infant born OOH | Case study of very premature birth |
|  | **Schramm et al (36)** | 1987 | **Neonatal mortality in Missouri Home Births, 1978-84** | USA | To review planned and unplanned home births and describe neonatal outcomes and mortality by birth attendant type, compared to physician attended hospital births during the same period. | Birth parents who had planned and unplanned home births in Missouri, USA | Retrospective analysis of Missouri planned and unplanned home births from 1978-1984. Compared to physician attended hospital births. |
|  | **Schultz et al (64)** | 2020 | **Epidemiology of oxytocin administration in out-of-hospital births attended by paramedics** | Australia | To describe the epidemiology and adverse side effects of oxytocin administration during OOHbs. | PCRs for OOHbs from QAS. | Retrospective analysis was undertaken of all OOHbs between the 1<sup>st</sup> January 2018 and 31<sup>st</sup> December 2018 attended by the Queensland Ambulance Service. |
|  | Scott et al (61) | 2005 | Unplanned Out of Hospital Births - Who Delivers the Babies? | UK | To review the incidence of OOHBs from South Tyneside District General Hospital and evaluate the birth attendants and time of day for these events. | PCRs for all OOHBs | Retrospective analysis of OOHBs occurring between 011099 and 310902 |
|  | Thornton & Dahlen (46) | 2018 | Born before arrival in NSW, Australia (2000–2011): a linked population data study of incidence, location, associated factors and maternal and neonatal outcomes. | Australia | To describe the incidence of BBAs in New South Wales (NSW), location, associated factors, and outcomes. | Four government reports: Perinatal Data Collection (PDC); Admitted Patient Data Collection; Australian Bureau of Statistics Mortality Data (ABS); NSW Registry of Births, Deaths and Marriages (RBDM). | A retrospective linked population data study involving population-based surveillance systems was undertaken for the years 2000–2011 (OOHBs compared to hospital births) |
|  | Unterscheider et al (31) | 2011 | Born before arrival births: Impact of a changing obstetric population. | United Kingdom (Ireland) | To describe the incidence of BBAs in the Rotunda Hospital, Dublin, Ireland, outcomes and possible associated factors. | PCRs from McKesson hospital database | Retrospective observational. OOHBs compared to in hospital births. |
|  | Verdile et al (20) | 1995 | Out-of-hospital deliveries: A five-year experience | USA | To review all OOHBs which occurred in ALS EMS care from 1984-1988 to determine the frequency and outcomes of these cases. | All PCRs from OOHBs attended by ALS EMS clinicians identified from the City of Pittsburgh, Bureau of EMS database | Retrospective observational review of five-years of OOHBs in EMS care |
|  | Wyckoff & Periman (34) | 2009 | Effective ventilation and temperature control are vital to outborn resuscitation | USA | To review the characteristics of OOH neonates, review early clinical management and perinatal outcomes compared to neonates born in-hospital. | PCRs from OOH neonates in the Parkland Memorial Hospital database, between January 1994 and December 2000. | Retrospective case-control study over a seven-year period of OOHBs |
| Resuscitative hysterotomies | Authors | Year of publication | Title of article | Country | Study aims | Participants | Methods |
|  | Gatti et al (26) | 2014 | Out-of-hospital perimortem cesarean section as resuscitative hysterotomy in maternal posttraumatic cardiac arrest | Italy | Case study of a traumatic maternal out-of-hospital cardiac arrest (OHCA), with resuscitative hysterotomy | One traumatic OHCA with perimortem cesarean section | Case study of resuscitate hysterotomy |
|  | Kupas et al (27) | 2009 | Case Conference: Out-of-hospital perimortem cesarean section | USA | Case study of a perimortem cesarean section | One OHCA with perimortem cesarean section | Case study of perimortem cesarean |
|  | Lenz et al (24) | 2017 | Out-of-hospital perimortem cesarean delivery performed in a woman at 32 weeks of gestation: A case report | Norway | Case study of a perimortem cesarean section | A birth parent at 32 weeks gestation, who experienced cardiac arrest | Case study of perimortem cesarean |

| Patient or paramedic experiences articles | Authors | Year of publication | Title of article | Country | Study aims | Participants | Methods |
| --- | --- | --- | --- | --- | --- | --- | --- |
|  | Dietsch et al (72) | 2010 | "Mind you, there's no anaesthetist on the road": women's experiences of labouring en route. | Australia | To explore birth parent's experiences of being in labour while being transported to a centralised maternity service, in NSW Australia. | 12 birth parents who were in labour and one partner | Qualitative interviews |
|  | Erlandsson et al (70) | 2015 | Women's experience of unplanned out-of-hospital birth in Sweden – a phenomenological description | Sweden | Aim was to capture birth parent's experience of unplanned OOH in Sweden. | Women who birthed OOH between 2010 and 2012 in Sweden. | Qualitative interviews using a phenomenological approach |
|  | Flanagan et al (71) | 2019 | Listening to women's voices: the experience of giving birth with paramedic care in Queensland, Australia. | Australia | To explore maternal perspectives on accessing care, and how previous birthing experiences and healthcare interactions influenced decisions on when to access care | Women who had experienced an unplanned OOH | Narrative inquiry, underpinned from a feminist perspective (interviews) |
|  | Flanagan et al (73) | 2019 | Women's experience of unplanned out-of-hospital birth in paramedic care | Australia | To explore the experiences of 22 women who birthed in paramedic care. | Women who birthed in paramedic care in Queensland, Australia between 2011 – 2016 | Narrative enquiry interviews. |
|  | Foster & Maillardet (56) | 2012 | Baby on the way: Was an ambulance in the plan? | United Kingdom | To survey the views of women who had requested an ambulance due to imminent birth. | All imminent birth patients during a four consecutive month period from August to November 2008 | Qualitative questionnaire |
| Duplicate record- appears under epidemiology and patient/clinician perspectives | Goodwin et al (65) | 2022 | Temperature measurement of babies born in the pre-hospital setting: analysis of ambulance service data and qualitative interviews with paramedics. | UK | To examine the proportion of instances where neonatal temperature is documented in PCRs by paramedics attending OOHs. Explore barriers and facilitators for paramedics in undertaking a temperature measurement of neonates. | PCRs for OOH neonates from the South West Ambulance Service | Two phase mixed method study. Phase I = retrospective observational examination of deidentified PCRs from 1 Feb 2017 to 31 Jan 2020. Phase II = purposive sampling; semi structured qualitative interviews via telephone with paramedics in Jan 2021 |
|  | Hill et al (8) | 2022 | Paramedic training, experience, and confidence with out-of-hospital childbirth (OOHB) in Australia | Australia | To investigate paramedic perceptions of training, experience, and confidence with OOH | Australian registered paramedics | Semi-structured qualitative interviews |
|  | Jarneid et al (68) | 2020 | Fathers' Experiences of Being Present at an Unplanned Out-of-Hospital Birth: A Qualitative Study. | Norway | To explore father's experiences at being present during an unplanned OOH. | Fathers present during an unplanned OOH | Qualitative semi-structured interviews conducted for fathers present at an unplanned OOHs occurring between 2015-2020 |
|  | **Persson et al (57)** | 2019 | **Specialist ambulance nurses' experiences of births before arrival.** | Sweden | To describe BBA experiences from specialist ambulance nurses, from two ambulance stations in Sweden. | Nurses with a minimum of 2-years' experience and who were qualified as specialist ambulance nurses. | A qualitative study was conducted, and nine specialist ambulance nurses who had assisted with one or more prehospital births were interviewed. |
|  | **Svedberg et al (6)** | 2020 | **Women's experiences of unplanned pre-hospital births: A pilot study** | Sweden | To describe women's experiences of OOHBs in ambulance nurse's care, in Sweden. | Women who had experienced an OOHB and requested an ambulance. | A qualitative questionnaire consisting of six open-ended questions that encouraged participants to describe their prehospital-birth experience. |
|  | **Vagle et al (58)** | 2019 | **Emergency medical technicians' experiences with unplanned births outside institutions: A qualitative interview study** | Norway | To investigate EMTs' experiences regarding OOHBs in Western Norway | EMTs from Western Norway who had experience with OOHB | Individual semi-structured interviews with 12 emergency medical technicians in Norway. |
|  | **Vik et al (69)** | 2016 | **Women's experiences with giving birth before arrival** | Norway | To describe women's experiences of OOHBs in Western Norway. | Birth parents who had experienced an unplanned OOHB | Semi-structured qualitative interviews. |

**Supplementary Table 2:** Identified risk factors for out-of-hospital births and increased risk for neonatal morbidity and mortality.

| Identified risk factors | Risk Factors for OOHb (or patients transferred to hospital) | Increased risk for neonatal morbidity and mortality | Proportional or descriptive Citations | Uni or Multivariate Regression Citations |
| --- | --- | --- | --- | --- |
| Alcohol use | x |  | (3,30) |  |
| Antenatal care – none or inadequate | x | x | (3,31–36) | (5,37–40) |
| Apgar <7 at 5-minutes |  | x | (41) | (42) |
| Bacterial vaginosis | x |  | (43) |  |
| Birth parent not originally from the country being studied. | x |  | (30,36) |  |
| Birth occurred between 1700 hours - midnight | x |  | (41) |  |
| Birth occurred between midnight - 0800 hours | x |  | (36,41) | (17) |
| Bleeding (active) | x |  | (43) | (44) |
| Breech presentation | x | x | (41) | (5) |
| Concealed or unknown pregnancies | x |  | (30,41) |  |
| Diabetes (type 1 or 2) | x |  | (33,43) |  |
| Did not experience rupturing of membranes prior to labour commencing | x |  | (45) |  |
| Dispatch request came between 0700 and 1859 hours | x |  |  | (44) |
| Dispatch request more common in spring and summer | x |  |  | (44) |
| Distance considered adjacent to the birthing unit/hospital | x |  | (45) |  |
| Distance >35 km/ travel time >45 mins to birthing unit | x |  |  | (30,37–39,46) |
| Ethnicity |  | x | (30,31,33,35,39) |  |
| Extreme prematurity (<28 weeks gestation) |  | x |  | (42) |
| Gestational age - lower compared to inborn or non-transferred neonates | x | x | (41) | (42) |
| Gestational diabetes | x |  | (3) |  |
| Gestational hypertension | x |  | (33,43) |  |
| Group B streptococcus | x |  | (43) |  |
| HIV positive | x |  |  | (40) |
| Hypertension | x |  | (3) |  |
| Illicit drug use | x | x | (3,30,31,33,43) |  |
| ‘Infectious disease’ | x |  | (30) |  |
| Intercurrent physician/midwife consultation |  | x |  | (5) |
| Less Basic Life Support provided during COVID lockdown | x |  |  | (17) |
| Living alone | x |  | (38) |  |
| Location of birth (during COVID lockdown- at home more) | x |  |  | (17) |
| Location of call request (urban/close to hospital) | x |  | (32) | (44) |
| Low / disadvantaged socioeconomic background | x |  | (30,34) | (46) |
| Maternal age (older) | x | x | (3,35,36,44,47) |  |
| Maternal age (younger) | x | x | (3,35,36,41) | (40,42,44) |
| ‘Maternal pathology’ |  | x |  | (5) |
| Meconium-stained fluids |  | x |  | (5) |
| Midwife consultation < 24 hours (or hospital consult) | x |  | (36) | (5) |
| Miscarriage (history) | x |  |  | (44) |
| Multigravida or multiparity | x | x | (3,30–32,36,41,45) | (5,37,38,44,46) |
| Neonate birth weight (low) |  | x | (31,36) | (40,42,46) |
| Neonatal death within 12 months |  | x | (31,41) | (40,42,46) |
| Neonatal hypothermia |  | x | (31) | (5,29,40) |
| Neonate intubation <24 hours |  | x |  | (40) |
| NICU admission | x | x | (31,36,41) | (46) |
| Not booked in to birthing suite | x |  | (30,32,41) |  |
| Not made contact with hospital prior to labour commencing | x |  | (45) |  |
| Obesity (or overweight) | x |  | (3,36,48) |  |
| Pelvic inflammatory disease | x |  | (43) |  |
| Prematurity (<37 weeks gestation) |  | x |  | (5,46) |
| Previous caesarean section | x |  | (43,48) |  |
| Previous short second stage of labour | x |  | (45) |  |
| Previous out-of-hospital birth | x |  | (32) |  |
| Primiparity | x | x | (3) | (5,44) |
| Prolonged preterm rupture of membranes (PPROM) | x |  |  | (40) |
| Short labours | x |  | (32,36,45) | (37) |
| Simple analgesia provided adequate pain relief | x |  | (45) |  |
| Single parent | x |  | (30,35,41) | (37) |
| Singleton pregnancy |  | x |  | (42) |
| Smoking | x | x | (3,30,35) | (37,46) |
| Stillborn |  | x | (42) | (42) |
| Stillbirth following antepartum haemorrhage |  | x |  | (42) |
| Trauma | x |  | (43) |  |
| Twins or multiple birth | x |  | (33,43) |  |
| Unemployed / unskilled / poor education | x | x | (30,35,41) | (38) |
| 'Unhealthy house' |  | x |  | (5) |
| Urinary tract infection | x |  | (43) |  |

**Supplementary Table 3:** Peripartum complications encountered by emergency ambulance clinicians.

| Complications | Citation |
| --- | --- |
| Abruption placentae | (23) |
| Amnionitis | (49) |
| Amniotic pulmonary embolism | (25,26) |
| Antepartum haemorrhage | (3,16,23,25,42–44,49–52) |
| Assistance required for third stage of labour | (36,45,49) |
| Chills | (48) |
| Concealed or unknown pregnancy | (18,19,23,30,37,38,41) |
| Cramping and/or pain | (3,8,16,23,30,34,43,44,50,52–58) |
| Eclampsia | (16,49,50) |
| Ectopic or molar pregnancy | (16,52) |
| En caul | (20) |
| Endometritis | (48) |
| Extended neonatal hospitalisation | (47) |
| Failure of descent | (3,59) |
| Foetal acidosis | (49) |
| Foetal bradycardia / non-reassuring foetal heart rate / abnormal foetal heart rate / distress | (23,43,48,49,59) |
| Foetal death | (22,25,54) |
| Foetal tachycardia | (49) |
| Gestational diabetes | (2,3,50,52) |
| ‘High’ birth weight / large neonate | (36,37,46) |
| Jaundice | (37,47) |
| ‘Low’ Apgar | (3,5,24,26,37,41,42,55,60) |
| ‘Low’ birth weight | (19,23,31–33,35–40,42,46,52,60,61) |
| Malposition (breech) | (2,3,5,16,20,23,34,40,41,48,49,52,54,58,60,62–64) |
| Malposition (face or unidentified) | (2,21,22,49,59) |
| Malposition (shoulder dystocia) | (2,5,21,23,46,58,64) |
| Maternal altered mental state | (52) |
| Maternal asthma | (2,50) |
| Maternal cardiac arrest and/or death (including resuscitative hysterotomy) | (16,21,23–27) |
| Maternal comorbidities | (52,64) |
| Maternal diabetes (type 1 or 2) | (2,33,43,46,50) |
| Maternal drug or alcohol use (illicit or prescription) | (2,3,16,21,30,31,33,37,43,50) |
| Maternal epilepsy | (2,50) |
| Maternal faintness, dizziness, weakness | (16,27,52) |
| Maternal fever | (3,52) |
| Maternal headache | (52,64) |
| Maternal Hepatitis C | (2) |
| Maternal HIV+ | (40) |
| Maternal hypertensive conditions | (2,3,16,20,33,43,46,49,50) |
| Maternal hypotensive conditions | (19,33,43,44) |
| Maternal hypoglycaemia | (3) |
| Maternal multiple sclerosis | (2) |
| Maternal psychiatric conditions | (2,50) |
| Maternal respiratory distress | (21,52) |
| Maternal seizure | (49,52) |
| Maternal suspected infection | (30,43,52) |
| Maternal tachypnoea | (2) |
| Maternal tachycardia | (19,43,44,53) |
| Maternal trauma | (21,25,26,43,50,52) |
| Meconium-stained fluid | (5,23,36,49,59) |
| Multiple birth (or multiple pregnancy) | (2,3,16,19,20,22,30,33,34,40,43,46,49,58) |
| Nausea/vomiting | (16,43,48,52,64) |
| Necrotising enterocolitis | (40) |
| Neonatal 'birth trauma' | (21,41) |
| Neonatal bradycardia | (3) |
| Neonatal bradypnoea | (3) |
| Neonatal congenital abnormality | (22,35,46,52) |
| Neonatal cyanosis | (20) |
| Neonatal death | (2,8,23,27,28,30–32,34,35,38–42,46,49,53,54,60) |
| Neonatal fractured clavicle | (48) |
| Neonatal hypoglycaemia | (3,32,37,40,47) |
| Neonatal hyperthermia | (29,31,32,39) |
| Neonatal hypothermia | (3,5,24,29–32,37–40,45–47,61,65) |
| Neonatal jaundice | (47) |
| Neonatal meconium aspiration | (19) |
| Neonatal patent ductus arteriosus | (40) |
| Neonatal seizure | (48) |
| Neonatal rash | (55) |
| Neonatal resuscitation (CPR) | (3,8,19–22,24,25,28,41,45,55) |
| Neonatal respiratory resuscitation (and/or intubation) | (3,19–22,24,26,28,38,41,43,46–48,55) |
| Neonatal suspected infection | (38,40,46,48) |
| Neonatal tachypnoea | (32) |
| Neonatal tachycardia | (37) |
| Neonatal withdrawal syndrome/ abstinence syndrome | (30,31,46) |
| NICU admission | (27,28,30–33,36–38,41,45,46,61) |
| Nuchal cord | (16,20–23,64,66) |
| Parietal diastasis | (47) |
| Pelvic disproportion | (59) |
| Perineal or vaginal trauma / suturing | (19,22,30,32,36,41,45–48,55,59,61) |
| Placenta previa | (3,25) |
| Pneumothorax | (48) |
| polyhydramnios | (49) |
| Postpartum haemorrhage | (2,3,6,8,16,18–22,30,32,41,45–48,52,53,55,59,60,64,65) |
| Post-term pregnancy | (43,48,49) |
| Preeclampsia | (2,16,48–50) |
| Premature birth | (2,3,5,16,18,19,22,23,28,30,32–34,36–38,40–42,45,46,48,52,53,55,58,60,61,67) |
| Premature rupture of membranes (PROM) | (34,42,48,49,59) |
| Preterm prelabour rupture of membranes (PPROM) | (28,40,43) |
| Previous caesarean section | (59) |
| Prolonged first or second stage of labour / fatigue | (3,23,48,49,55,56,59) |
| Retained placenta | (3,30,41,45,47,55,59) |
| Sphincter rupture | (60) |
| Spontaneous or induced abortion | (16,19,22,52) |
| Stillbirth | (3,20,36,37,40,42,53,68) |
| Suspected uterine growth retardation | (49) |
| Threatened miscarriage | (3) |
| Threatened preterm labour | (16,28,43,44,49,50,61) |
| Threatened preterm labour with ruptured membranes | (28,43,44) |
| Umbilical cord prolapse / rupture | (2,3,16,49,64) |

**Supplementary Table 4.** Emergency ambulance clinician interventions and/or management for OOHBs.

| Intervention / management | Citation |
| --- | --- |
| Apgar score | (2,3,5,8,19,20,26,27,31,37,39,42,46,59,60,66) |
| Assist with birth | (2,3,5,8,18–22,28,30,32,35–39,41,43–45,47,49,50,52,55,56,61,63,64,66) |
| Attempted to assist with birth (unsuccessful) | (34,54,62) |
| Clamp/cut the cord | (2,21,39,58) |
| Fluids administration (including red blood cells) | (2,21,26,39,40,50,53) |
| Foetal doppler | (66) |
| Fundal massage (both appropriate and inappropriately applied) | (2,3,39) |
| Gynaecological examination | (43,44) |
| Haemorrhage control (ante partum or post partum) | (8,18,50,53,55,64,65) |
| Management of third stage (either physiological or active) | (2,3,18,39,41,53,61,64,66) |
| Maternal airway aspiration/suctioning | (21,24,26) |
| Maternal cardiac monitoring | (2,50,55,66) |
| Maternal IV (intravenous) cannulation | (2,21,24–26,39,44,50,53,55,56,62,66) |
| Maternal oxygen administration (+BVM, PPV) | (2,21,24,26,27,44,50,54,55,69) |
| Maternal positioning: left lateral tilt, Trendelenburg position, displacement of uterus | (25,55,66) |
| Maternal resuscitation cardiopulmonary (BLS and ALS, incl Lucas Device) | (23–27) |
| Maternal resuscitative hysterotomy | (23–27) |
| Maternal tracheal intubation | (24–26) |
| Medication administration (including adrenaline, amiodarone, antibiotics, atropine, diltiazem, dopamine, Entonox, fentanyl, glucose, heparin, insulin, labetalol, magnesium, magnesium sulphate, methoxyflurane, metoclopramide, midazolam, morphine, nitroglycerine, oxytocin, prochlorperazine, propofol, salbutamol, surfactant, ‘tocolytics’, thrombolysis, tranexamic acid) (maternal or neonatal) | (2,3,6,8,18,21,23–26,40,43,44,49,50,55–57,64,66) |
| Needle decompression | (26) |
| Neonatal hypothermia management | (3,33,37,65) |
| Neonatal hypoglycaemia management | (3,33) |
| Neonatal IO cannulation | (2,26,55) |
| Neonatal IV cannulation | (21,62) |
| Neonatal resuscitation (cardiopulmonary) | (2,3,17,19,20,23–25,27,30,33,41,55) |
| Neonatal resuscitation (respiratory) inclusive of airway clearance/suctioning, O2 | (2,3,8,20,21,23,24,26,27,33,37,39,41,55) |
| Neonatal surfactant administration | (24) |
| Neonatal tracheal intubation (or attempt) | (2,19,23,24,26,27,33,39,40,55) |
| Neonatal – temperature | (65) |
| Nuchal cord | (16,20,23,64,66) |
| Rupture membranes | (39) |
| Skin-to-skin care and other rewarming techniques (drying, bonnets, wrapping, ambient temperature control, incubator) | (29,39,58) |

**Supplementary Table 5:** Barriers and challenges for emergency ambulance clinicians when managing out-of-hospital births.

| Education and training | Communication and collaboration | Environmental | Technology and aids | Other |
| --- | --- | --- | --- | --- |
| *Emergency ambulance clinicians feel underprepared for these infrequent callouts -> desire more training. (3,8,19,37,54,57,58,60,65,69,74)<br>*Clinicians lack confidence with attending OOHs. (8,57,58,74)<br>*Authors or clinicians suggest further ongoing training re OOHs is necessary. (3,6,8,17,19,20,22,23,30,32,38,40,45,47,49,52,53,57,66,67,71)<br>*Specific skills training required in:<br>-oxygen administration for neonate. (21,33,40)<br>-third stage. (45)<br>....-hypothermia for neonate. (5,33,39,40,65)<br>-birthing shoulders for breech. (34,62)<br>*Early recognition of complications might not occur. (8)<br>*Desire for Obstetrics placements. (8,58)<br>*Pregnancy-specific guideline adherence is low. (25,26) | *Midwives 'gatekeeping' hospital admissions. (69,71)<br>*High level of coordination required for aeromedical retrievals. (66)<br>*Improved collaboration between organisations would be beneficial (ambulance/hospital/ maternity services/midwives). (36,46,50,52,53,55–58,69,74)<br>*Role and responsibility uncertainty when midwives present. (60,65)<br>*No debriefing provided to emergency ambulance clinicians after an OOH. (57,58) However debriefings were provided for some. (25,27)<br>*Language barriers with patients. (5,19,30,36,47,50,52,58,74)<br>*May overlook birth plan. (8,73)<br>*Communication with birth parent may become 'clinical' or not optimal, if highly focussed on situation. (8,69,73)<br>*Desire for midwives to accompany ambulance. (58,69)<br>*Not gaining consent. (73) | *Confined or complex birth location (ie: ambulance, toilet, in-flight etc). (2,3,5,6,17,19,20,22,26,32,36,37,41,47,52,53,56–58,60,61,63,66,68,69,72)<br>*Difficult extrication. (58,72)<br>*Flights subject to weather. (66)<br>*Aircraft / boat may require modifications for obstetrics cases. (66)<br>*Neonatal resuscitation difficult at home. (30,31,33)<br>*Distance or time to definitive care. (8,28,30,37–39,46,48,49,57,58,60,64,69,72,74)(21)<br>*Temperature regulation difficult especially for neonate (promote skin-to-skin). (5,19,29,37,39,40,45,60,66,69,70,72)<br>*Long on-scene time (maternal cardiac arrest). (25,50,55,58)<br>*Difficulty in locating patients already in transit, particularly in rural areas. (72) | *Limited ability to determine foetal wellbeing. (25,50)<br>*Incomplete Patient Care Record information, or not creating a second Patient Care Record for neonate. (2,3,16,21,22,30,38,39,50,53,65,74)<br>*Need appropriate neonatal thermometer. (3,65)<br>*Prompts required to ensure neonatal temperature is taken. (65)<br>*Mobile phone coverage. (8,69,72)<br>*No neonatal restraints in the ambulance. (57)<br>*Limited obstetrics equipment / medication available. (21,22,57,58)<br>*Desire for telehealth support. (8)<br>*Desire for prompts or memory aids, including electronically or manually. (21,29,65) | *Challenges of managing two patients. (16,57,65)<br>*Guidelines specific to OOH situations. (5,16,21,40,53,54,67,70,74)<br>*Lack of clinician experience apparent to patient and bystanders. (68,69,73)<br>*Increase in OOHs following SARS-CoV-2. (17)<br>*Lack of pain relief options. (8,72)<br>*Access to obstetrics services no longer close to home. (6,72,74)<br>*Managing labour / reducing contractions. (43)<br>*Bystanders can complicate scenes and patient care. (53)<br>*Refusing transport. (3,52,58)<br>*Patient insists on female clinician attending. (58)<br>*No/low antenatal care. (2,3,5,20,23,31–41,52,61)<br>*Birth before clinician arrival. (2,3,5,6,8,18–20,29,32,36,37,39–41,45,46,61,64,74)(22) |

## Notes

### Competing Interest Statement

The authors have declared no competing interest.

### Funding Statement

This research was undertaken with assistance from a PhD scholarship awarded as part of Dr Luke Hoppers ECU Vice Chancellors Research Fellow Scholarship for 2020. There are no grant/award numbers associated with this.

